# Anomalous epithelial variations and ectopic inflammatory response in chronic obstructive pulmonary disease

**DOI:** 10.1101/2020.12.03.20242412

**Authors:** Naoaki Watanabe, Jun Nakayama, Yu Fujita, Yutaro Mori, Tsukasa Kadota, Iwao Shimomura, Takashi Ohtsuka, Koji Okamoto, Jun Araya, Kazuyoshi Kuwano, Yusuke Yamamoto

## Abstract

Phenotypic alterations in the lung epithelium have been widely implicated in Chronic obstructive pulmonary disease (COPD) pathogenesis, but the precise mechanisms orchestrating this persistent inflammatory process remain unknown due to the complexity of lung parenchymal and mesenchymal architecture. To identify cell type-specific mechanisms and cell-cell interactions among the multiple lung resident cell types and inflammatory cells that contribute to COPD progression, we profiled 52,764 cells from lungs of COPD patients, non-COPD smokers, and never smokers using single-cell RNA sequencing technology. We predicted pseudotime of cell differentiation and cell-to-cell interaction networks in COPD. While epithelial components in never-smokers were relatively uniform, smoker groups represent extensive heterogeneity in epithelial cells, particularly in alveolar type 2 (AT2) clusters. Among AT2 cells, which are generally regarded as alveolar progenitors, we identified a unique subset that significantly increased in COPD patients, and specifically expressed a series of chemokines and PD-L1. A trajectory analysis revealed that the inflammatory AT2 cell subpopulation followed a unique differentiation path, and a prediction model of cell-to-cell interactions inferred significantly increased intercellular networks of inflammatory AT2 cells. Our results identify previously unidentified cell subsets and provide an insight into the biological and clinical characteristics of COPD pathogenesis.

## Introduction

Chronic obstructive pulmonary disease (COPD) is a chronic inflammatory disease mainly caused by long-term cigarette smoke (CS) exposure. It has emerged as the fourth cause of mortality globally ^1^. COPD is characterized by persistent airflow limitation, which is progressive even after smoking cessation ^2^. The inflammatory process of COPD is predominantly localized to the small airways and surrounding lung parenchyma with concomitant systemic inflammation ^3^. Phenotypic alterations in the lung epithelium caused by CS exposure have been widely implicated in COPD pathogenesis ^4^, but the precise mechanisms orchestrating this persistent inflammatory process remain unknown.

The distal airway system is composed of a variety of epithelial cells as well as immune and mesenchymal lineages. Among epithelial populations, alveolar type 1 (AT1) and AT2 cells constitute the alveolar epithelium. On the other hand, basal lineages such as club and ciliated cells reside in the bronchiolar epithelium. AT2 cells are multifunctional and polarized epithelial cells spatially restricted to distal lungs. AT2 cells functions as progenitors of AT1 cells, contributing to the maintenance of alveolar homeostasis ^5^. AT2 dysfunction has been implicated in the pathogenesis of a variety of parenchymal lung diseases, including COPD, acute respiratory distress syndrome, and idiopathic pulmonary fibrosis (IPF) ^4,6,7^. With respect to protective roles in COPD pathogenesis, club cells in bronchiolar epithelia are well-known to exert effects on anti-inflammation and immunomodulation ^8,9^. As such, the cellular complexity of lung hinders a deeper understanding of cell-specific roles of disease pathogenesis. Therefore, little is known about not only cell type-specific mechanisms but also cell-cell interactions among the multiple lung resident cell types and inflammatory cells that contribute to COPD progression.

The development of single-cell RNA sequencing (scRNA-seq) has provided a new research tool to obtain gene expression profiles at single-cell resolution and identify novel cellular phenotypes in lung diseases, such as IPF and asthma ^10–13^. Although some scRNA-seq studies have revealed the diversity of aberrant lung resident cell landscapes in COPD lungs ^12,14^, the cellular heterogeneity and cell type-specific mechanisms of COPD progression have not been fully investigated. To improve our understanding of the cell-specific mechanisms in COPD pathogenesis, we used scRNA-seq to reveal the mechanism of COPD pathogenesis and elucidate differences in the proportions and transcriptional phenotypes of epithelial cells in COPD and identified inflamed epithelial populations such as a unique AT2 cell subset. Analysis of intercellular communications revealed that this AT2 cell subset had the strongest interactions with immune cells in COPD patients. We generated novel insights into phenotypic changes in epithelial cells and altered communication patterns between the COPD-specific epithelial populations and immune cells.

## Results

### Single-cell composition in the lungs of patients with COPD

We initially performed single-cell transcriptome profiling from human lung tissues of 6 COPD, 3 non-COPD smoker, and 3 never-smoker samples (**Figure 1A**). The lung tissues were obtained from lobectomy specimens of primary lung cancer. Pathohistological observation clearly showed morphological differences between COPD, non-COPD smokers, and never-smokers (**Figure 1B** and **Supplemental Table 1, 2)**. In the process of quality control with the Seurat R package, the sequencing data of the estimated dead cells were computationally excluded, and to reduce batch effect of each sample, the data was integrated with “Harmony”^15,16^.

**Figure 1.**
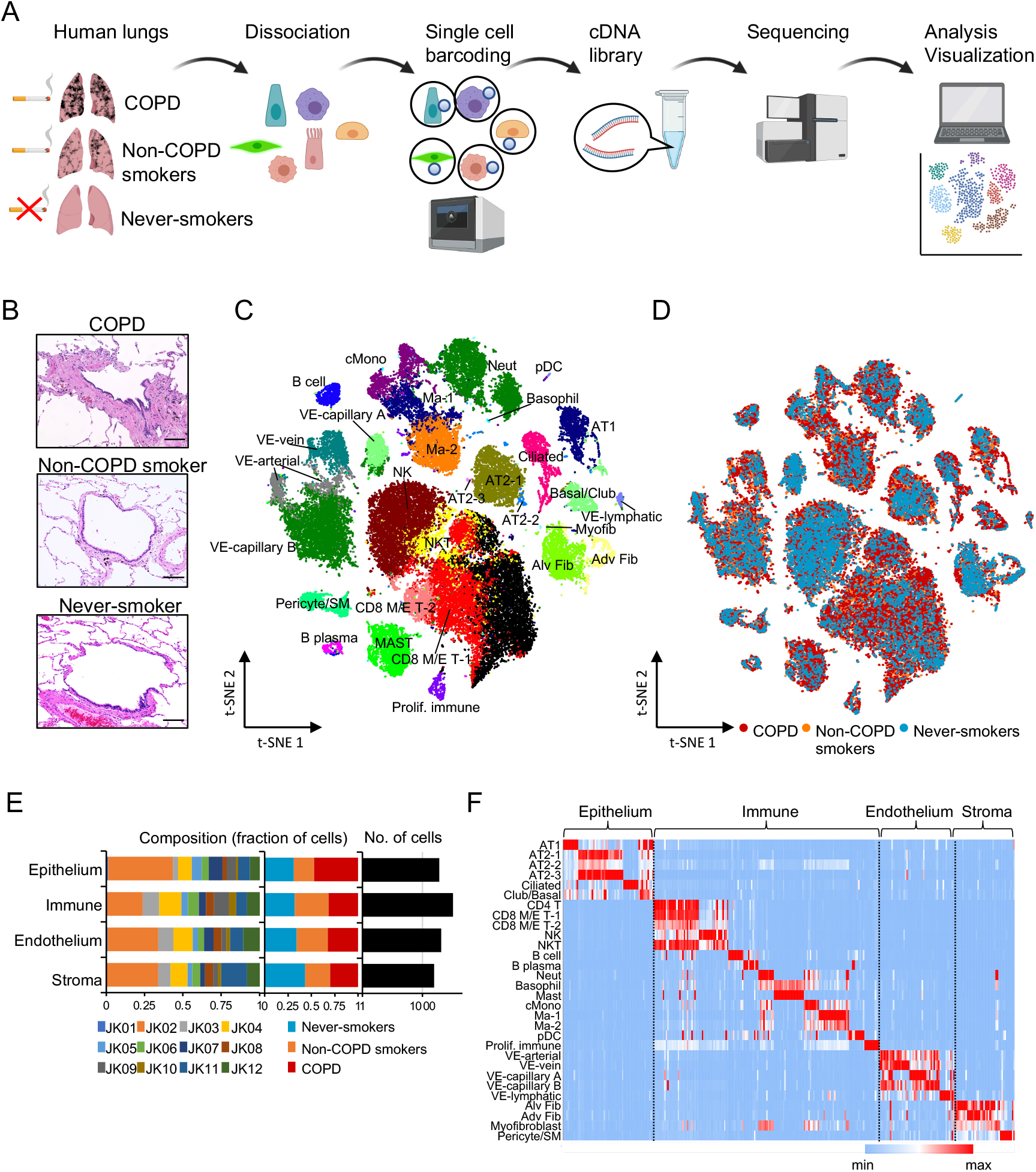
Charting the human lung cell landscape of COPD patients, non-COPD smokers and never-smokers by scRNA-seq. **A.** Experimental flowchart for scRNA-seq. Patient lung tissue samples were enzymatically digested into single cells. cDNA synthesis was performed using a 10x genomics Chromium controller system, and libraries were sequenced on Illumina platforms. **B.** Representative H&E images of COPD, non-COPD smoker, and never-smoker lung sections. **C.** t-SNE plot displaying the major clusters of human lung cells. **D.** t-SNE plot classifying patient states such as COPD, non-COPD smokers, and never-smokers. **E.** Fraction of cells (x-axis) from each sample (left) and patient state (middle) in each cluster from the t-SNE plot (y-axis). The cell numbers of each cluster are shown (right). **F.** Heatmaps of differentially expressed genes. The average expression (log2[TPM + 1]) of the top 10 genes (row) in each cluster (columns) is shown. NK, natural killer cell; NKT, natural killer T cell; Ma, macrophage; Neut, neutrophil; cMono, classical monocyte; pDC, plasmacytoid dendritic cell; VE, vascular endothelial; AT, alveolar type; CD4 T, CD4^+^ T cell; CD8 M/E T, CD8^+^ memory/effector T cell; Alv Fib, alveolar fibroblast; Adv Fib, adventitial fibroblast; Myofib, myofibroblast; Proif.immune, proliferating immune cell; SM, smooth muscle.

Unsupervised clustering followed by t-distributed stochastic neighbor embedding (t-SNE) revealed 30 distinct clusters (**Figure 1C** **and Supplemental Table 3**). A similar result was obtained by UMAP (**Supplemental Figure 1A**). Almost no difference between with and without batch effect cancellation by Harmony was observed in PCA (**Supplemental Figure1, B and C**). According to the gene expression profiles, this clustering analysis identified 15 clusters of immune-type cells, such as neutrophil and NK cells; 6 clusters of epithelial lineages, such as AT2 cells, AT1 cells and basal cells; and 4 clusters of stromal-type cells, such as fibroblasts, and smooth muscle cells and 5 clusters of endothelial cells (**Figure 1C**).

When remapping single cells with the patient states (COPD, non-COPD smokers, and never-smokers), although the variation in cell distribution was evident, all clusters contained cells from three patient states (**Figure 1D**). The number of cells in each cluster varied among patients and states, and most clusters were distinctly represented in each patient (**Figure 1E**). Clustering with t-SNE represents the differences in gene expression profiles in the clusters. We further investigated cluster-specific gene signatures and selected the top 10 specifically expressed genes in each of the clusters (**Figure 1F, Supplemental Figure 1D, and Supplemental Table 4**) ^17^. Thus, these data reveal that our single-cell transcriptome successfully represented the cellular clusters in COPD, non-COPD smokers, and never-smokers.

### Differential cellular distribution of the immune, endothelial, and stromal cell components in COPD lungs

Given the importance of immune cell populations in COPD pathogenesis, we initially focused on immune cells in patients with COPD. As illustrated in **Figure 1C**, our dataset comprised a number of immune cell-related clusters, representing 63.2% (33,365 cells) of human lung cells. To scrutinize the immune cell population, single-cell data of immune clusters (**Figure 1C**) were selected, and a UMAP plot with all immune cells was redeveloped (**Figure 2A**). Similar to the t-SNE mapping of all lung cells (**Figure 1D**), the UMAP plot of immune-related cells revealed the overall proportion of immune cells among the patient states (**Figure 2, B-D**). In 15 immune cell clusters, we found two discrete cell populations for macrophages and CD8^+^ memory/effector (M/E) T cells (**Figure 2, A and D**). For example, INHBA was highly expressed in macrophage-1 but not in macrophage-2; GPR183 was specifically expressed in macrophage-2, although macrophage receptors with collagenous structures (MARCOs) were highly expressed in both clusters (**Figure 2E**). The heatmaps displayed distinct expression patterns of cluster-specific gene signatures (**Figure 2F**). Comparative analysis of cell numbers in the clusters among the patient states revealed that NK cell and CD8 M/E T cell-1 numbers were significantly altered in COPD and non-COPD smoker samples (**Figure 2G**). In the clustering of scRNA-seq datasets, we could not distinguish Treg cells from the CD4^+^ T cell cluster; however, the UMAP plot with FOXP3 expression strikingly showed a small population of Tregs in the CD4^+^ T cell cluster (**Supplemental Figure 2A**). To evaluate the reliability of our dataset, we integrated several public available datasets (**see Methods**). The UMAP plots of lymphocytes and myeloid cells in combined publicly-available datasets showed distinct molecular clusters similar to ours (**Figure 2H**). We found that resistin (RETN) which has been reported to have a function in inflammation and immune responses and to act as a pro-inflammatory mediator was highly expressed in COPD alveolar macrophage ^18^. We confirmed this finding with combined publicly-available datasets (**Figure 2I**).

**Figure 2.**
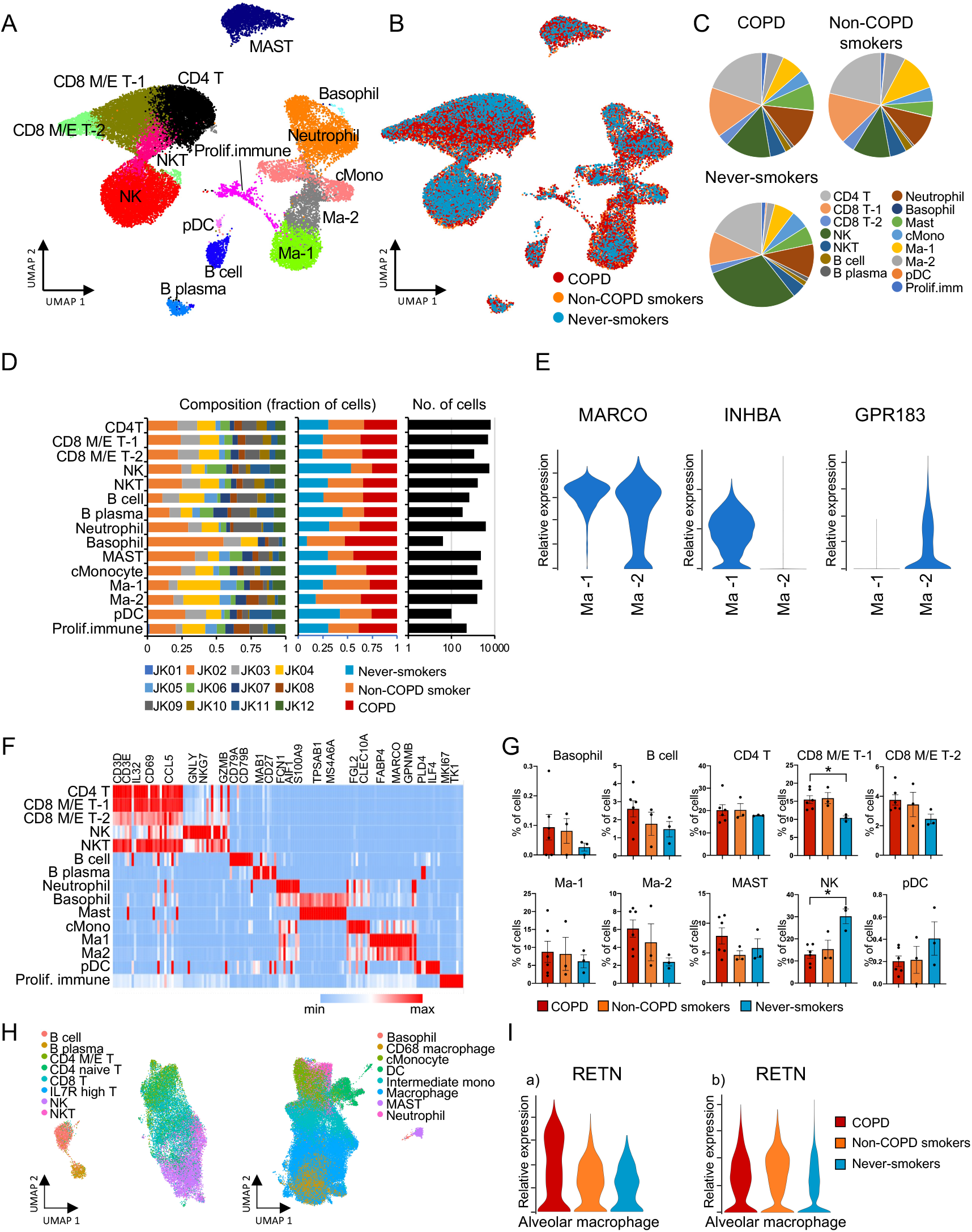
Single-cell transcriptome analysis of the immune components of human distal lung tissues from COPD patients, non-COPD smokers, and never-smokers. **A.** a UMAP plot displaying the major immune cell clusters of human lung cells. **B.** a UMAP plot of immune components classifying the patient states, such as COPD, non-COPD smokers, and never-smokers. **C.** Pie charts displaying immune cell compositions among COPD, non-COPD smokers, and never-smokers. **D.** Fraction of cells (x-axis) of immune components from each sample (left) and patient state (middle) in each immune cluster (y- axis). The cell numbers of each cluster of immune components are shown (right). **E.** Violin plots for typical markers and differentially expressed genes among macrophage clusters. **F.** Heatmap of differentially expressed genes across immune cell clusters. The average expression (log2[TPM + 1]) of the top 10 genes (row) in each immune cluster (columns) is shown. **G.** Bar charts displaying percentages of subpopulations for immune cell clusters across the patient states. p < 0.05 by Tukey’s test (COPD vs. never-smokers). **H.** UMAP plots of lymphocytes (left) and myeloid (right) cells in combined publicly- available datasets. **I.** Violin plots for RETN in alveolar macrophages. a) present dataset; b) combined publicly-available datasets.

Next, we sought to investigate endothelial and stromal cell populations in COPD lungs. The t-SNE plot of all lung cells contained five endothelial clusters and four stromal cell clusters, representing 16.2% (8,522 cells) and 13.5% (7,139 cells), respectively (**Figure 1D**). For further analysis of endothelial and stromal cells at single-cell resolution, these clusters of all lung cells were selected, and a UMAP plot with endothelial and stromal cells was redeveloped in the same manner as the immune cell analysis.

In the UMAP plot of endothelial and stromal cells, we found three distinct cell populations in fibroblasts (**Figure 3, A and D**). All fibroblast clusters expressed general fibroblast markers such as DCN and PDGFRA. A difference in the expression levels of a small number of genes defined the fibroblast clusters. For example, FGFR4 was highly expressed in alveolar fibroblast, and PDGFRL was specifically expressed in adventitial fibroblast (**Figure 3E**). The heatmaps displayed distinct expression patterns of cluster-specific gene signatures (**Figure 3F**). Comparative analysis of cell numbers in the clusters among patient states revealed that VE-capillary A was significantly decreased in COPD samples; on the other hand, the cell numbers in the VE-vein cluster was increased in COPD and non-COPD smoker samples (**Figure 3G**). IL6 is known to be highly expressed in COPD lungs ^19^; our scRNA-seq revealed that it was particularly upregulated in adventitial fibroblasts (**Figure 3I**). Furthermore, the UMAP plots of stromal and endothelial cells in combined publicly-available datasets are shown in **Figure 3H**. We found that ASAP1, involved in the regulation of cytoskeleton remodeling, was upregulated in COPD alveolar fibroblast (**Figure 3Ja**) in our data sets and also confirmed this finding with combined publicly-available datasets (**Figure 3Jb**) ^20^.

**Figure 3.**
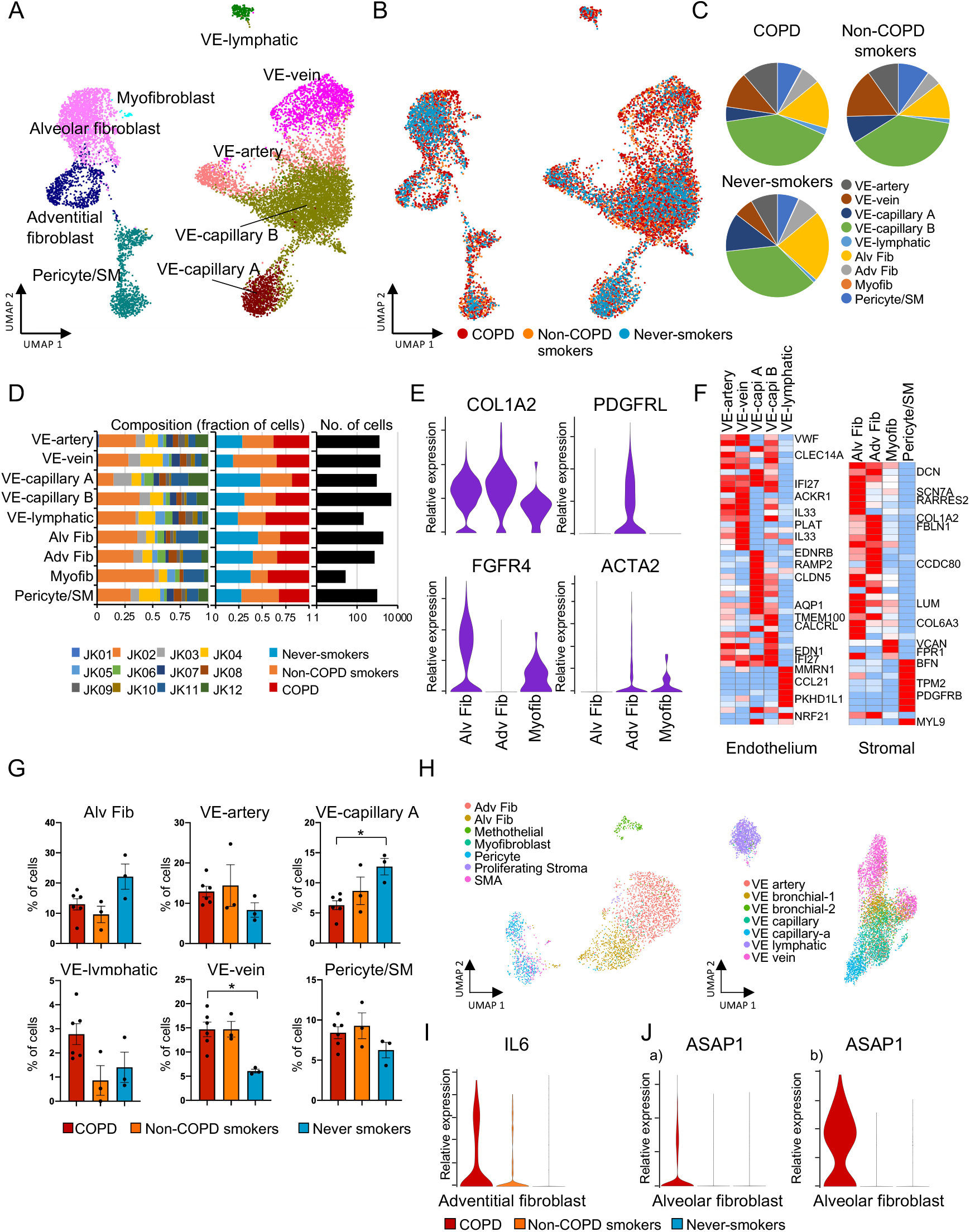
Single-cell transcriptome analysis of the endothelial and stromal components of human distal lung tissues among COPD, non-COPD smokers, and never-smokers. **A.** a UMAP plot displaying the major endothelial and stromal clusters of human lung cells. **B.** a UMAP plot of endothelial and stromal components classifying patient states such as COPD, non-COPD smokers, and never-smokers. **C.** Pie charts displaying endothelial and stromal cell compositions among COPD, non-COPD smokers, and never-smokers. **D.** Fraction of cells (x-axis) of endothelial and stromal components from each sample (left) and patient state (middle) in each stromal cluster (y-axis). The cell numbers of each cluster of stromal components are shown (right). **E.** Violin plots for typical markers and differentially expressed genes among fibroblast clusters. **F.** Heatmap of differentially expressed genes across endothelial and stromal clusters. The average expression (log2[TPM + 1]) of the top 10 genes (row) in each endothelial and stromal cluster (columns) is shown. **G**. Bar charts displaying percentages of subpopulations for endothelial and stromal cell clusters across the patient states. p < 0.05 by Tukey’s test (COPD vs. never-smokers). **H.** UMAP plots of stromal (left) and endothelial (right) cells in combined publicly-available datasets. **I.** Violin plots for IL6 in adventitial fibroblast. **J.** Violin plots for ASAP1 in alveolar fibroblast. a) present dataset; b) combined publicly- available datasets.

Our scRNA-seq analysis of immune, stromal, and endothelial cells apparently showed cellular states in COPD, non-COPD smokers, and never-smokers and found a difference in cell populations between the COPD and never-smoker samples. We also identified specific gene profiles of COPD in alveolar macrophage and alveolar fibroblast.

### Precise and extensive classification of epithelial lineages in COPD

To investigate epithelial heterogeneity in COPD, a total of 6,823 epithelial cells were re-clustered (**Figure 4, A and B**). The UMAP plot with epithelial lineages revealed 10 clusters, including two AT1 cell clusters, three AT2 cell clusters, and two ciliated cell clusters. Few neuroendocrine cells ^21^ were found in the cluster of club cells (**Supplemental Figure 2B**). Presumably due to too few cells, they did not form a unique cluster. We did not find ionocyte ^22^ and tuft ^23^ clusters (**Supplemental Figure 2, C and D**). Analysis of closeness centrality in epithelial cells revealed a decreased centrality in the COPD epithelial cells than in the non-COPD smoker and never-smoker cells (**Figure 4C**), suggesting increased diversity in the COPD epithelia. The density UMAP plot showed that a large number of cells were present in the AT2 and AT1 clusters (**Supplemental Figure 2E**). As expected, the cell numbers of AT2 and AT1 cells in COPD samples tended to decrease compared with those in non-COPD samples (**Supplemental Figure 2F** and **Supplemental Table 5**). Cell populations among the patient states are summarized in **Figure 4, D and E**, which exhibit remarkable differences in epithelial cell composition.

**Figure 4.**
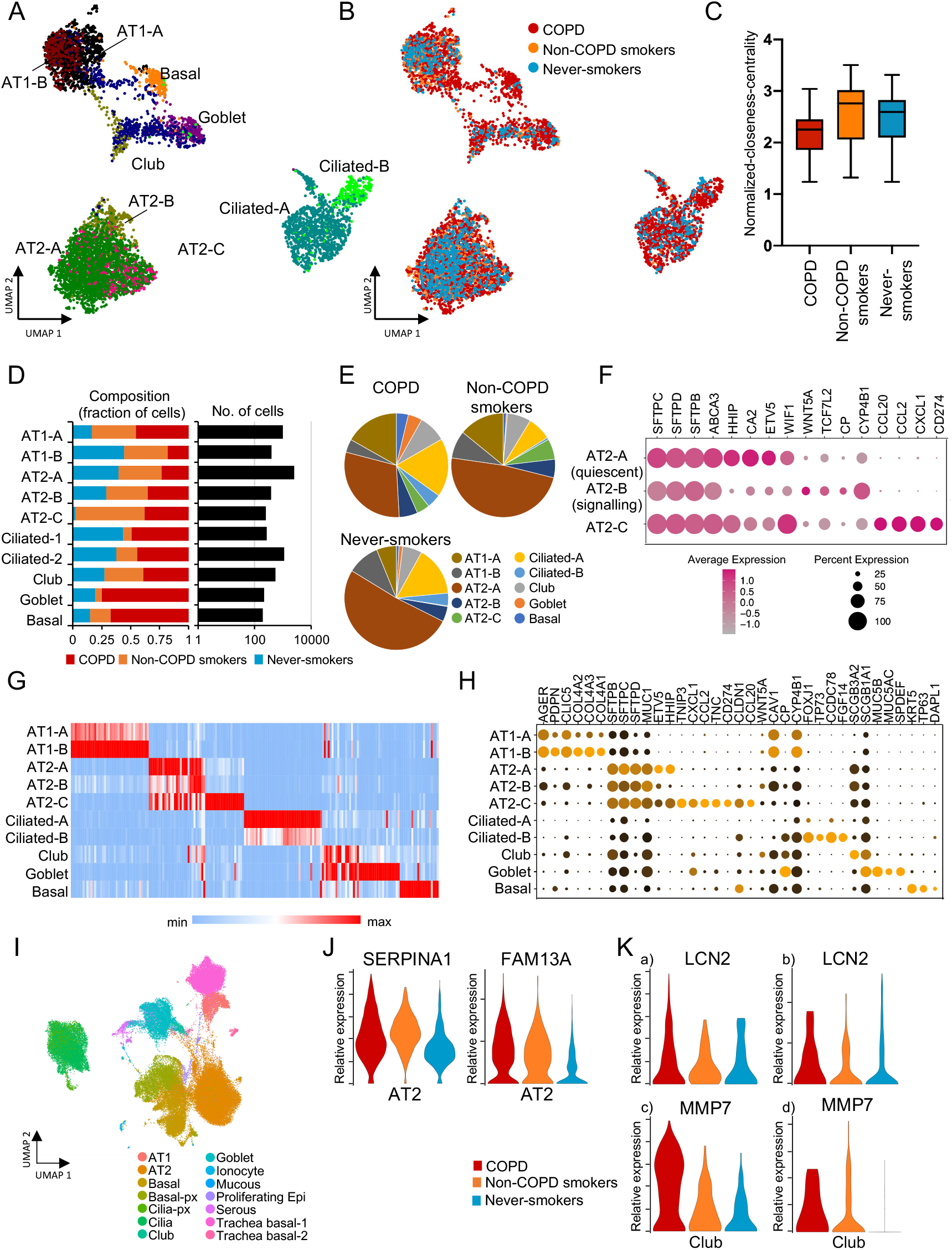
Single-cell transcriptome analysis of the epithelial components of human distal lung tissues among COPD, non-COPD smokers, and never-smokers. **A.** a UMAP plot displaying the major epithelial clusters of human lung cells. **B.** a UMAP plot of epithelial components classifying patient states such as COPD, non-COPD smokers, and never-smokers. **C.** Box plot of normalized closeness centrality across COPD, non-COPD smokers, and never-smokers. **D.** Fraction of cells (x-axis) from the patient state (left) in each cluster from the UMAP plot (y-axis). The cell numbers of each cluster of epithelial components are shown (right). **E.** Pie charts displaying epithelial cell compositions among COPD, non-COPD smokers, and never-smokers. **F**. Heatmap showing the average expression levels and percentage of expression of selected marker genes in the three AT2 clusters. **G.** Heatmap of differentially expressed genes across epithelial clusters. The average expression (log2[TPM + 1]) of the top 20 genes (row) in each epithelial cluster (columns) is shown. **H.** Heatmap showing the average expression levels and percentage of expression of selected marker genes in the epithelial cluster. **I.** a UMAP plot of epithelial cells in combined publicly-available datasets. **J.** Violin plots for SERPINA1 and FAM13A in AT2 cells. **K.** Violin plots for LCN2 in club cells. a) present dataset; b) combined publicly-available datasets. Violin plots for MMP7 in club cells. c) present dataset; d) combined publicly-available datasets.

We identified three epithelial cell clusters of AT2 cells. SFTPC, a typical marker for AT2 cells, was strongly expressed in all clusters; however, subtle variations in gene expression among the AT2 clusters were observed (**Figure 4F**). For example, Hedgehog interacting protein (HHIP) and inhibitors of Wnt (WIF1) signaling were highly expressed in AT2-A, presumably implicating that they are quiescent AT2. In contrast, WNT5A and detoxification gene, CP were expressed in AT2-B cluster, suggesting that these cells could be alveolar progenitors, AT2-signalling (AT2-s) ^24,25^. These data suggest epithelial cell diversity in human lung cells. The differentially expressed genes in each of the clusters are summarized (**Figure 4, G and H**) and exhibit the diversity of epithelial lineages. Differential gene markers among cell-types were shown in separated heatmaps (**Supplemental Figure 2G**).

In addition, we found particular expressions of FAM13A and SERPINA1 in AT2 cells (**Figure 4J**), as reported to be highly expressed in COPD lungs ^26^. The UMAP plot of epithelial cells in combined publicly-available datasets is shown in **Figure 4I**. We identified an increased expression level of the innate immunomodulator lipocalin-2 (LCN2) in the COPD club cells ^27^. Matrix metallopeptidase 7 (MMP7), which functions to facilitate the repair and regulate the acute inflammatory response, was upregulated in the COPD club cells (**Figure 4K**). Furthermore, we found upregulation of FK506 binding protein 5 (FKBP5), a known negative regulator of the glucocorticoid receptor that directly regulates corticosteroid anti-inflammatory function ^28^, in not only epithelial but also most the cells including immune and stromal cells (**Supplemental Figure 4C**).

### Unique epithelial cell populations and an inflammatory AT2 cluster in the COPD lungs

We next explored the unique or increased cell populations in the COPD lung epithelia. The cell populations of the AT1 subtypes, AT2 subtypes, and basal lineages were separately compared among COPD, non-COPD smoker, and never-smoker samples (**Figure 5A**). In the AT1 subtypes, the AT1-A cluster is the major cell cluster in COPD (80.9% in AT1 subtypes of COPD); however, this cluster represents a small population in never-smokers (39.5% in the AT1 subtypes of never-smokers). In contrast, the AT1-B clusters, which specifically expressed COL4A1, COL4A2, and COL4A3 (**Supplemental Figure 4B**), decreased in the COPD (never-smokers: 60.5%; non-COPD smokers: 46.0%; COPD: 19.1%) (**Figure 5A, top panel and Supplemental Figure 4B**). In the AT2 subtypes, the cell population of the AT2-A cluster obviously decreased in the COPD (**Figure 5A, middle panel**). The AT2-C clusters appeared in the smokers (COPD: 25.1%; non-COPD smokers: 30.7%; never-smokers: 0.32%). In basal lineages, the cell populations of goblet (MUC5B, MUC5A, and SPDEF), club (SCGB3A1, CYP2F2, and CCKAR), and basal cell (KRT17, KRT5, and TP63) clusters were higher in COPD than in never-smokers (**Figure 5A, bottom panel**). Furthermore, we found that the percentage of the subpopulations in AT1-B, AT2-A, and AT2-B were lower in the COPD than in the others (**Figure 5B, upper panels**). We also observed that the percentage of the club, goblet, basal cell, and AT2-C clusters tended to be high in the COPD (**Figure 5B, lower panels, and** **Figure 5C**). The difference becomes more apparent when comparing COPD and others (non-COPD smokers and never-smokers) (**Supplemental Figure 2H**). Because the cells located in the AT2-C cluster were hardly detected in the never-smokers, we further scrutinized the cell population and characterized the gene expression profile of the AT2-C cluster. As shown in **Figure 5C**, the percentage of AT2-C cluster cells in the total cell numbers of lung tissues was very low (0.45% in COPD). Notably, a number of inflammation-related genes, including CXCL1, CXCL8, CCL2, CCL20, and CD274 (PD-L1) were specifically expressed in the AT2-C cluster, and we designated the population inflammatory AT2 (iAT2) (**Figure 4F**, **Figure 5D, and Supplemental Figure 3A**). In our combined publicly-available datasets, we confirmed the existence of a population with a high expression of chemokines, including CXCL2 and CXCL8 in the COPD AT2 cells (**Figure 4E**).

**Figure 5.**
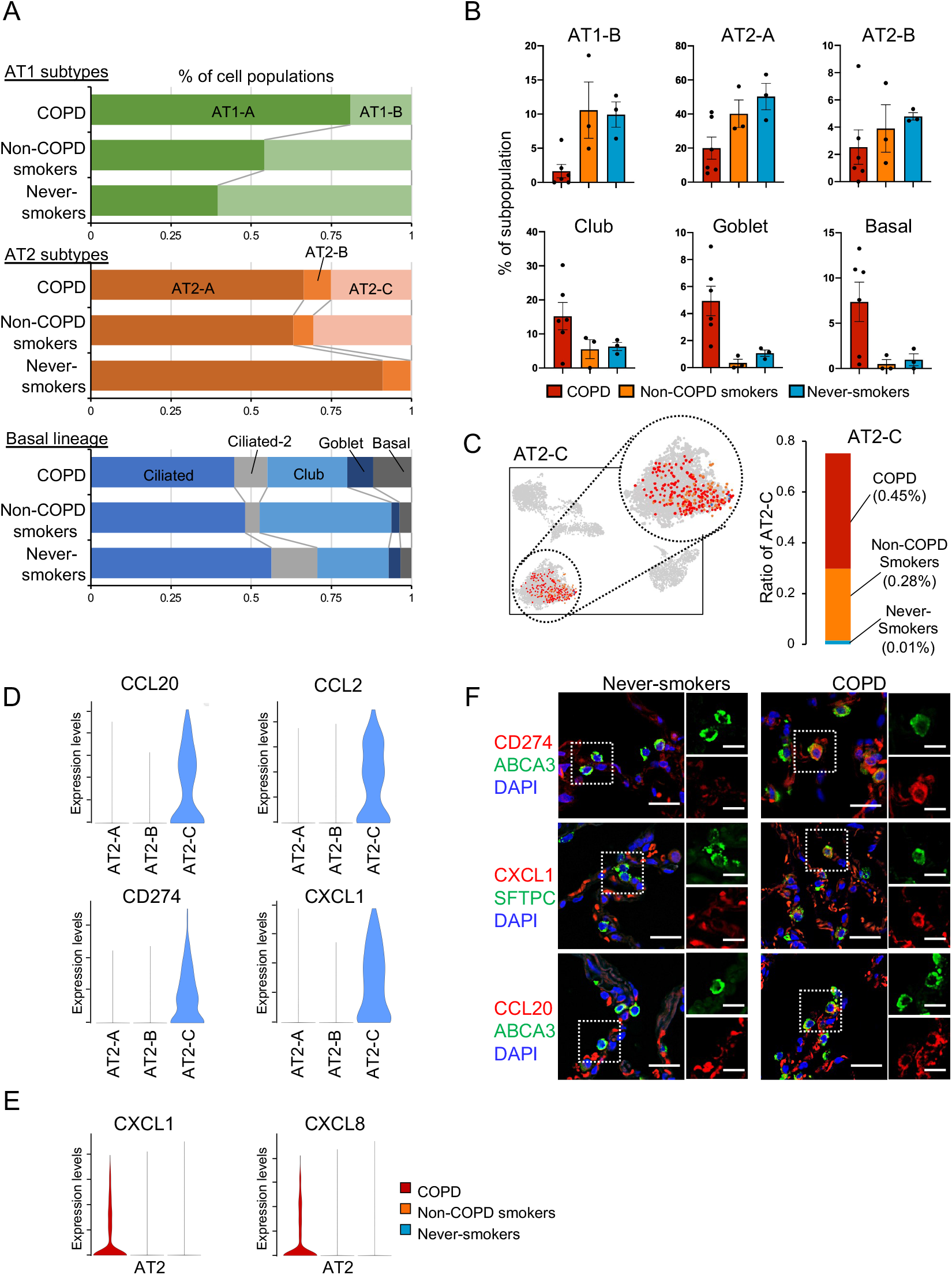
Variations in epithelial cell components among COPD, non-COPD smokers, and never-smokers. **A.** 100% stacked bar charts of the percentage of cell populations for AT1 subtype clusters, AT2 subtype clusters, and basal lineage clusters. **B.** Bar charts displaying the percentages of subpopulations such as AT1-B, AT2-A, AT2-B, club, goblet, and basal clusters across patient states. **C.** a UMAP plot of epithelial cells focusing on the AT2-C cluster (left). The cell population of the AT2-C cluster (right). The y-axis represents the ratio of cells in the AT2-C cluster across patient states. Brackets () represent the percentage of cells of the AT2-C cluster based on epithelial cells in each of the patient states. **D.** Violin plots of representative inflammatory-related genes displaying the AT2-C (iAT2) cluster. **E.** Violin plots for CXCL1 and CXCL8 in AT2 cells in combined publicly-available datasets. **F.** Representative images immunostained with CD274 (PD-L1), CXCL1 and CXCL20 with AT2 cell markers (SFTPC or ABCA3) in COPD and never-smoker lung sections. Scale bars, 25 μm (left), 12.5 μm (right).

Additionally, we identified novel markers to classify the iAT2 cluster, such as CLDN1 and TNC (**Supplemental Figure 3B**). The specific expression of the inflammatory genes and the markers that we identified were confirmed with immunofluorescence (**Figure 5F**). CCL20 was not detected in the AT2 cells of the never-smokers; however, very few ABCA3-positive AT2 cells in COPD coexpressed CCL20. Likewise, we found coexpression of CXCL1 and SFTPC, CD274 and ABCA3, CCL2 and SFTPC, TNC and ABCA3, and CXCL8 and SFTPC in the COPD lung alveolar tissues (**Figure 5F** **and Supplemental Figure 4A**).

### Distinct differentiation program and increased cellular variation in the COPD epithelia

To investigate how the iAT2 cluster appears in the COPD lungs, we sought to dissect the inferred differentiation trajectories of epithelial lineages. For this purpose, we performed pseudotime analyses ^29^ for alveolar (**Figure 6, A-C**) and basal lineages (**Supplemental Figure 5, A-B**), separately. Because AT2 cells generally serve as the progenitors of AT1 cells, the plot for alveolar differentiation revealed a trajectory from AT2 to AT1 clusters (**Figure 6A, right panel**). A three-dimensional pseudotime plot clearly revealed that the iAT2 cluster located at the branches from the middle of AT2 cells, suggesting that iAT2 exhibited a distinct differentiation program (**Figure 6C**). In basal lineages, we observed bidirectional differentiation into either ciliated cell clusters or secretory cell clusters in all groups (**Supplemental Figure 5A**), although the cell composition differed among the disease states, e.g., increased populations of secretory and basal cells in the COPD (**Supplemental Figure 5B and** **Figure 5B, lower panels**).

**Figure 6.**
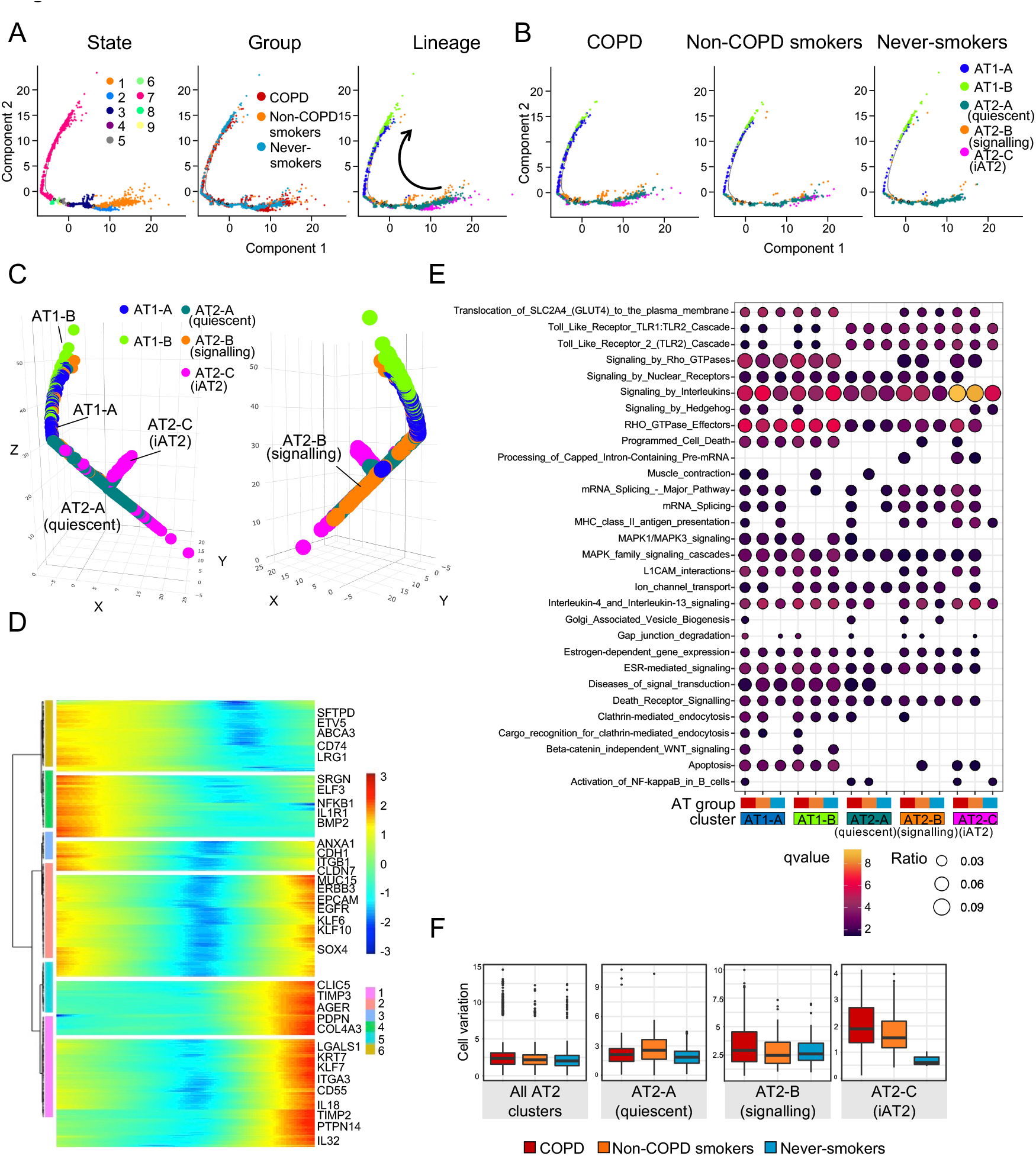
Pseudotime prediction of alveolar differentiation and epithelial variations in COPD. **A.** Pseudotime developmental trajectory analysis of alveolar lineages. Left panel: state, middle panel: group (COPD, non-COPD smokers, and never-smokers), right panel: alveolar clusters. Arrow: prediction of differentiation flow. **B.** Pseudotime plots for COPD, non-COPD smokers, and never-smokers. **C.** Three-dimensional pseudotime plots of alveolar lineages. **D.** Expression heatmap showing differentially expressed genes between two transition states (FDR < 0.01). Selected genes are highlighted. **E.** Pathway enrichment analysis of alveolar clusters among COPD, non-COPD smokers, and never-smokers. Color scale represents FDR q value. Dot size represents the likelihood ratio. **F.** Box plots showing the variation of clusters in COPD, non-COPD smokers, and never-smokers. Statistics are shown in **Supplemental Table 12**.

To understand the biological processes contributing to the pseudotime components, we analyzed gene expression that cooperatively changed with pseudotime. From the marker genes selected by the Seurat program, significantly altered gene groups (746 genes for alveolar clusters and 895 genes for basal clusters, false discovery rate (FDR) < 1%) were identified in the processes of alveolar and basal lineage differentiation (**Figure 6D** **and Supplemental Figure 5C**). In line with the distribution of the clusters, the AT2 cell markers, such as ETV5, were expressed in the predicted early phase, while the CLIC5 and AGER, AT1 cell markers, were highly detected in the late phase (**Figure 6D**). Likewise, we observed that basal cell markers, such as TP63, were expressed early and that ciliated cell markers, such as FOXJ1 and the DNAH family members, were expressed late (**Supplemental Figure 5C**).

We further investigated cluster-specific gene signatures by performing pathway enrichment analysis based on the Reactome database ^30^ for alveolar and basal lineages (**Figure 6E** **and Supplemental Figure 5D**). In **Figure 6E**, we found clear differences in the pathway enrichment between the AT1 and AT2 clusters. While two AT1 clusters exhibited relatively uniform patterns of pathway enrichment, more heterogeneity was observed within the AT2 clusters. In addition, diversity in pathway enrichment was also found among the patient states. For example, in the iAT2 clusters, “L1 CAM interactions” were enriched in COPD and non-COPD smokers but not in never-smokers (**Figure 6E**). Next, we calculated cellular variation among three AT2 clusters and showed a large variation in COPD (**Figure 6F**). iAT2 exhibited the largest variation in COPD, suggesting that COPD pathogenesis increased epithelial cell diversity in AT2 cells.

### Anomalous cell-to-cell networks in COPD epithelia

As COPD epithelial cells exhibited increased variations in expression profiles and ectopic inflammatory expression, we next explored receptor–ligand pairing between epithelial cells and immune cells. To this end, we focused on immunological responses, extracted chemokines and inflammatory cytokines from the Cell-Cell Interaction Database, and established an immune-response network between epithelial and immune cells. The interaction scores were calculated by the expression levels of receptors and ligands and the positive ratio in each cluster (**Methods**). We identified 981 predicted interactions (289 pairs between epithelial ligands and immune receptors and 692 pairs between immune ligands and epithelial receptors) in COPD (**Figure 7A**). We also analyzed the receptor– ligand pairing of non-COPD smokers and never-smokers in the same manner (**Supplemental Figure 6, A and B**). Among the epithelial clusters, the iAT2 cluster exerted the strongest effect on immune cells based on the interaction score (**Figure 7A**); the quantification of effect in COPD patients was remarkably higher than that in non-COPD smokers and never-smokers (**Figure 7, B-D**). While many interactions were commonly detected in all disease states, some were specific or strong to the COPD (**Figure 7D** **and Supplemental Table 6-11**).

**Figure 7.**
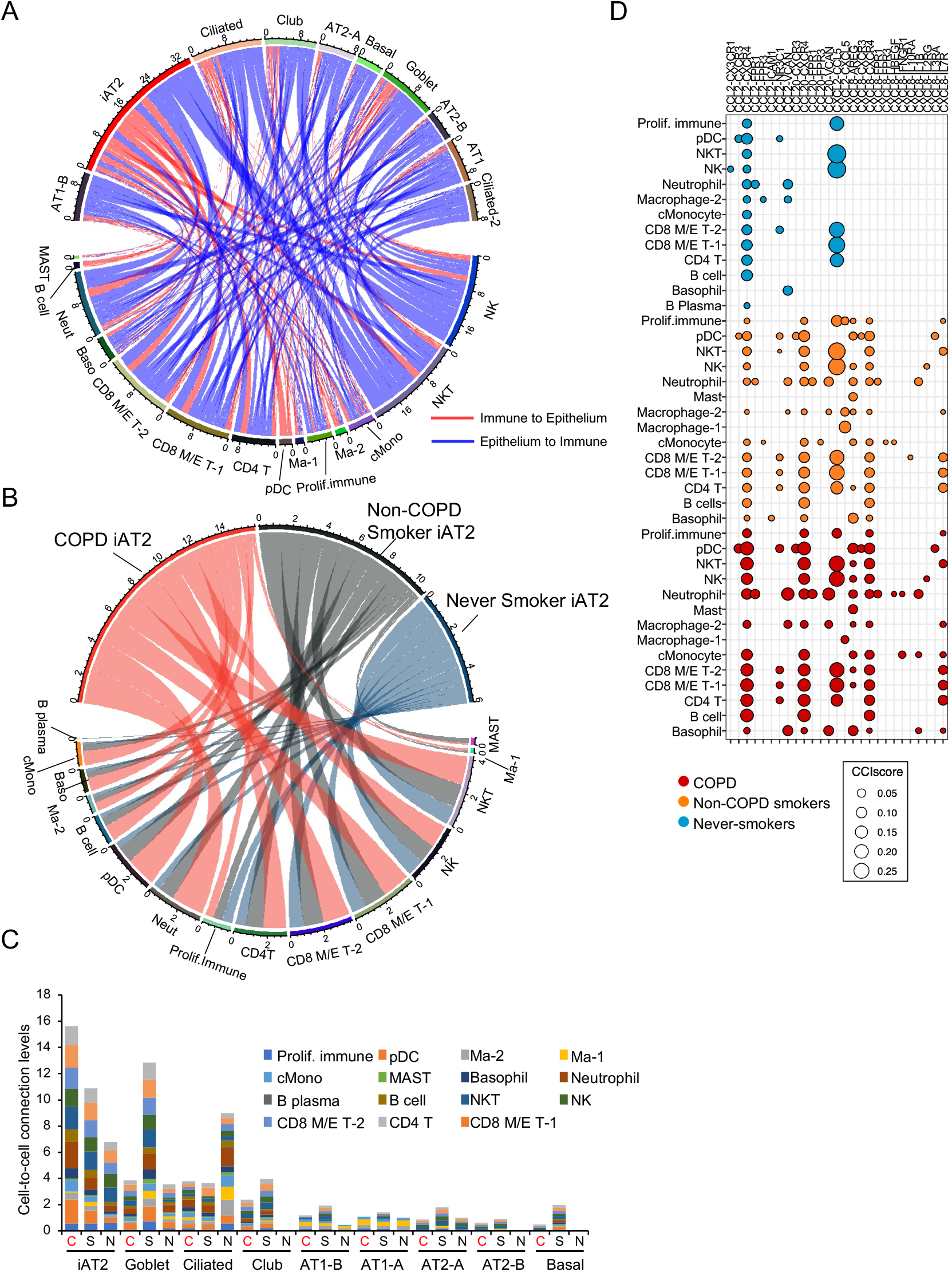
Inferred cell-to-cell interaction networks in the COPD epithelia. **A.** a Circos plot of cell-to-cell interactions featuring inflammatory cytokines and chemokines in COPD. Red lines represent predicted immune cell-ligand to epithelial cell-receptor interactions. Blue lines represent predicted epithelial cell-ligand to immune cell-receptor interactions. Ma, macrophage; Neut, neutrophil; cMono, classical monocyte; pDC, plasmacytoid dendritic cell. **B.** Ligand-receptor interactions on iAT2 clusters among COPD, non-COPD smokers, and never-smokers. **C.** Stacked bar charts showing cell-to-cell connection levels of epithelial cell-ligand to immune cell-receptor interactions. C: COPD, and S: non-COPD smokers, N: never-smokers. **D.** Bubble chart displaying cell-to-cell interactions of iAT2 clusters with immune cells. Red: COPD, orange: non-COPD smokers, and blue: never-smokers. Circle size represents the sum of the CCI score.

## Discussion

Chronic inflammation has been widely implicated in airway remodeling and emphysema development during COPD pathogenesis ^31–33^. This comprehensive single-cell transcriptome profiling of human lung tissues elucidates the presence of unique epithelial cell populations, implicating the inflammatory properties in COPD lungs. We identified 30 clusters of resident cell landscapes, including immune cells, epithelial cells, stromal cells, and endothelial cells. Based on further extensive classification, differential cellular distributions of the immune, endothelial, and stromal cell components were detected among the patient groups. Among immune-type cells, a significant decrease in NK cells was demonstrated in COPD and non-COPD smoker lungs. Although significant differences in cell numbers were observed in some clusters between smokers and never smokers, immune, endothelial, and stromal-type cells failed to show a clear difference in cellular distribution between COPD and non-COPD smoker lungs. It indicates that those alterations can be mainly attributed to CS exposure but may not directly reflect COPD pathogenesis. Extensive classification of epithelial lineages identified 10 clusters and elucidated not only the diversity of epithelial lineages but also marked differences in cell composition between COPD and non-COPD smoker lungs. In AT1 clusters, COL4A2 and COL4A3 were predominantly expressed in AT1-B cluster, and their expression levels gradually decreased along with COPD progression. Among the AT2 clusters, iAT2 exhibited remarkable variations in COPD by means of pathway enrichment analysis, suggesting the participation of AT2 diversity in COPD pathogenesis. The physiological participation of iAT2 cells in COPD pathogenesis was implicated by cell type-specific markers, including CCL20. In the basal lineage, increased populations of some secretory clusters were detected specifically in COPD samples. These goblet and club cell clusters also showed bidirectional interactions with immune cells. Taken together, these inflamed populations of the airway and alveolar epithelial cells could be involved in the mechanisms for the development of COPD lesions by exaggerating the inflammatory process.

Although COPD is the representative inflammatory lung disease induced by CS inhalation, the exact mechanism for persistent activation of lung inflammatory pathways even after smoking cessation remains elusive ^2^. Heterogeneity in airway inflammatory patterns has also been demonstrated in COPD lungs, which contain neutrophils associated with inflammasome activation, Th1- and Th17-mediated immunity, and Th2-mediated immunity^31^. During airway remodeling in COPD development, the airway epithelium undergoes profound changes in the numbers, proportion of cell types, and differentiation profiles ^32,33^. Furthermore, phenotypic alterations to cellular senescence in both the airway and alveoli have been implicated in COPD pathogenesis through excessive cytokine production of the senescence-associated secretory phenotype (SASP) ^34–37^. Accordingly, heterogeneity in epithelial phenotypic alterations with respect to inflammation may have a pivotal role in regulating not only CS-mediated transient responses but also the persistent nature of inflammatory processes in COPD lungs. Our comprehensive scRNA-seq analysis, especially focusing on epithelial phenotypic alterations, uncovered the large variation in gene expression profiles in COPD lungs (**Figure 4C** **and** **Figure 6F**) and the existence of inflamed airway and alveolar epithelial cell clusters, which are capable of mediating inflammatory processes through bidirectional interactions with immune cells (**Figure 7, A-D**). The cell-cell interactions and chemokine expression may partly explain the role of epithelial cells in CS-mediated activation of inflammatory signaling with a concomitant increased recruitment of neutrophils, CD4^+^ and CD8^+^ T cells into the lungs^2^. We obtained lung samples from lung cancer patients who had all ceased smoking for at least two months before the surgery to ensure a safe operation. Hence, it is likely that those inflamed epithelial clusters do not reflect the simple effect of CS exposure but rather intrinsic changes in phenotype.

AT2 has been recognized to play an essential role in maintaining alveolar homeostasis by not only secreting surfactant but also behaving as a tissue-resident progenitor cell for AT1 repopulation in response to lung injury. The pathogenic implication of alveolar epithelial cells in COPD development is not entirely clear, and the cells have been recognized to be passive sufferers of repetitive smoking stress with respect to cumulative cell loss mainly via regulated cell death ^38–40^. Furthermore, exaggerated loss of alveolar epithelial cells cannot be compensated for by an increase in proliferation of AT2 in COPD lungs, which is attributed to loss of stemness through enhanced cellular senescence induced by repetitive smoking stress ^35^, resulting in lung tissue destruction from emphysema development. Although recent advances in scRNA-seq technology have uncovered potential heterogeneity of the AT2 population^41^, we identified iAT2 cells with increased chemokine expression. Also, iAT2 cells showed increased expression of NFKB2, a component of NF-κB, and CSF3, an IL-6 superfamily member, further supporting the inflammatory nature of AT2 (**Supplemental Figure 3**). In contrast, the fraction of AT2 (quiescent state) clusters without an inflammatory phenotype was significantly decreased in COPD patients but occupied a major part of the AT2 population in never smokers, indicating that AT2 (quiescent state) is involved in a physiological role in normal lungs. The physiological involvement of iAT2 cell-mediated inflammation in the loss of parenchymal structure may be further strengthened by a recent report, which showed that the existence of β1 integrin-deficient AT2 with upregulation of NF-κB-dependent chemokines was sufficient for developing accelerated inflammation ^42^. A recent paper suggested the existence of a novel subpopulation of AT2 with high PD-L1 but low SFTPC expression in IPF ^43^. Despite the common characteristic of high PD-L1 expression, we recognize substantial differences between our iAT2 and AT2 subpopulations. Although the AT2 subpopulation with high PD-L1 but low SFTPC expression was responsible for the process of lung regeneration in IPF pathogenesis, we identified that iAT2 cells clearly exhibited an inflammatory phenotype associated with the potential loss of the lung structure of the COPD phenotype. However, the physiological role of high PD-L1 expression in iAT2 cells remains uncertain in terms of regulating cell survival, T cell function, and potential association with smoking-related lung cancer development.

There are a couple of limitations to our present study. One is the sample size. To clarify the existence of novel epithelial clusters with inflammatory phenotypes, we obtained data sets from 12 samples from a single center. Because it may not be sufficient for comprehensively evaluating COPD pathogenesis with high clinical heterogeneity, we confirmed our finding with combined publicly-available datasets of COPD. Another is an uncertain relationship between iAT2 function and COPD pathogenesis. Chronic CS exposure is a major cause of COPD, but approximately 20% of smokers develop clinically diagnosable COPD. The number of iAT2 cells was increased in both COPD (0.45%) and non-COPD (0.28%) lungs; hence, it is not deniable that iAT2 cells can exhibit a simple phenotypic alteration to AT2 cluster cells in response to chronic CS exposure. The pathogenic role of the iAT2 cell-mediated inflammatory process as a distinctive AT2 subpopulation in COPD pathogenesis remains unclear. The clinical implications of alterations in the basal lineage also remain elusive, especially in terms of airway remodeling and eosinophilic inflammation. The clinical implications of inflammatory epithelial cells should be examined using appropriate mouse models in future studies.

Overall, our comprehensive single-cell transcriptome profiling of COPD lung tissues identified cellular diversity and cell-specific characteristic gene expression in COPD. Notably, we identified inflammatory epithelial cell clusters which may explain the mechanisms for persistent activation of inflammatory pathways after smoking cessation. This finding implicates the possibility of orchestrating inflammation among a variety of cell types, resulting in airway remodeling and emphysema development in COPD pathogenesis.

## Methods

### Patient recruitment

Human lung tissue from never-smokers, non-COPD smokers, and patients with COPD was obtained from patients undergoing lobectomy for focal lung cancer. The diagnosis of COPD was established using the Global Initiative for Chronic Obstructive Lung Disease criteria. Informed consent was obtained from all surgical participants as part of an approved ongoing research protocol by the ethical committee of The Jikei University School of Medicine (#20-153 (5443)). Written informed consent was received by the patients.

### Lung tissue resection

Freshly resected lung tissue was intraoperatively obtained from patients with focal lung tumors, and normal and COPD parts of lung tissues were obtained from uninvolved regions. Isolated lung tissue fragments were mechanically minced and dissociated with enzymes according to a Lung Dissociation Kit protocol (130-095-927, Miltenyi Biotec, Bergisch Gladbach, Germany). After the dissociation, viability of single cells was confirmed more than > 90%, by trypan blue staining. Cells were then washed in phosphate-buffered saline (PBS) supplemented with 0.1% bovine serum albumin (BSA, Sigma-Aldrich, MO, USA), passed through 70-μM and 40-μM filters, centrifuged at 300 g for 10 minutes and resuspended in PBS supplemented with 1% BSA.

### Chromium 10x Genomics library preparation and sequencing

Cells were counted and resuspended in PBS supplemented with 0.1% BSA for loading for single-cell library construction on the 10x Genomics platform (10x Genomics, CA, USA). Experiments up to 4th samples were performed with the Chromium Single Cell 3′ GEM, Library & Gel Bead Kit v3 and Chromium Single Cell B Chip Kit according to the manufacturer’s instructions in the Chromium Single Cell 3′ Reagents Kits V3 User Guide. Experiments after the 5th sample were performed with the Chromium Next GEM Single Cell 3′ GEM, Library & Gel Bead Kit v3.1 and Chromium Next GEM Chip G Single Cell Kit according to the manufacturer’s instructions in the Chromium Next GEM Single Cell 3′ Reagents Kits V3.1 User Guide. Briefly, 6,000 cells were loaded into each channel and then partitioned into gel beads in emulsion in the Chromium Controller, where cell lysis and barcoded reverse transcription of RNA occurred, followed by amplification, fragmentation, and 5′ adaptor and sample index attachment. Sequencing was performed on a HiSeq X (Illumina, CA, USA) at a median sequencing depth of 112,482 reads/cell.

### Single-cell RNA sequencing data alignment

Raw sequencing data were processed with the Cell Ranger pipeline (version 3.0.2, 10x Genomics) and mapped to the human genome (GRCh38) to generate matrices of gene counts by cell barcodes.

### Data quality control and normalization

scRNA-seq analyses, including normalization, scaling, clustering of cells, and identifying cluster marker genes, were performed using the R software package Seurat version 3.2.^44^. We extracted single cells with nFeature_RNA > 500 and percent.mt < 20 for doublet and dead cell removal. Log normalization by function ‘NormalizeData’ from Seurat. Samples were log normalized and scaled for the number of genes and percentage of mitochondrial reads.

### Data clustering and dimensionality reduction

We performed principal component analysis (PCA) for dimensionality reduction in R with Seurat. To minimize the difference in batch effects, scRNA-seq data was integrated with Harmony^15,16^. Clustering of a single cell was performed by the functions ‘FindNeighbors’ and ‘FindClusters’ from Seurat. The dimensionality-reduced cell clustering is shown as a tSNE plot by the function ‘runTSNE’ and a UMAP plot by the function ‘runUMAP’. Clusters were grouped based on expression of tissue compartment markers (for example, EPCAM for epithelial cells, CLDN5 for endothelial cells, COL1A2 for stromal cells and PTPRC for immune cells) and then annotated in detail according to “A molecular cell atlas of the human lung” ^17^.

### Trajectory analysis

We used the ‘monocle’ package for trajectory analysis of alveolar cells and bronchiolar cells ^29^. Dimensionality reduction in the trajectory plot was performed by the ‘DDRTree’ method with the top 10% genes of the identified cluster markers.

### GO and pathway enrichment analyses

Pathway datasets were downloaded from the Reactome database. We performed enrichment analysis against the signature gene list from each cell cluster by the ‘ClusterProfiler’ and ‘ReactomePA’ packages in R ^30,45^. For enrichment analysis, gene symbols were converted to ENTREZ IDs using the ‘org.Hs.eg.db’ package (Carison M, R package version 3.10.0., 2019). GO enrichment analysis using the ‘enrichGO’ function and pathway enrichment analysis using the ‘enrichPathway’ function were performed by the BH method.

### Network analysis

Correlational network analysis was performed by the ‘igraph’ package as previously described ^46^. We calculated correlational coefficients between epithelial single cells in R, where r is the absolute value of Pearson’s correlational coefficient and n is the cell number of the cluster.

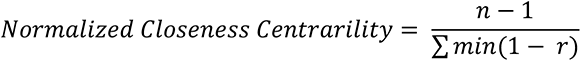

Network visualization was performed by ‘Y Organic Edge Router Layout’ in yfile programs using Cytoscape v3.7.1.^47,48^.

### Ligand-receptor cell-cell interaction (CCI) analysis

Ligand-receptor interactions between epithelial and immune/stromal cell subpopulations were performed using the interaction database of Bader’s laboratory from Toronto University (https://baderlab.org/CellCellInteractions#Download_Data) in R software. We selected the genes that were categorized as ‘interferons’, ‘interleukins’ and ‘TNFSF superfamily’ in the HUGO Gene Nomenclature Committee (https://www.genenames.org/). We calculated the cell number of subpopulations that expressed a value greater than 2. Only subpopulations whose expression cell ratio exceeded 10% were extracted for CCI network analysis, and the CCI score was calculated between each subpopulation. Next, a circos plot was drawn using the ‘circlize’ package in R ^49^. The circle plot used the sum of the CCI scores from subpopulation A to subpopulation B for all combinations.

L: Ligand subpopulation (ligand gene expression > 2), R: receptor subpopulation (receptor gene expression > 2), n: cell number.

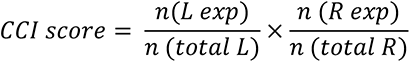

### Reanalysis of the public scRNA-seq datasets

To confirm the inflammatory signatures in COPD epithelial cells, we performed the reanalysis of scRNA-seq of COPD patients from the public cohort ^12^. The normalized scRNA-seq dataset was downloaded from Gene Expression Omnibus (GEO) and analyzed with the annotation metadata from GSE136831. It was analyzed and visualized by UMAP method with Seurat in R.

### Data availability

Transcriptomic data of scRNA-seq was deposited to the NCBI GEO database under accession number GSE173896.

### Code availability

The source code of scRNA-seq analysis is available on GitHub (https://github.com/JunNakayama/scRNAseq-COPD).

### Immunofluorescence

Each lung was fixed in 4% paraformaldehyde overnight and embedded in paraffin. Lung sections were dewaxed with xylene and rehydrated with ethanol (100–70%). Antigen retrieval was performed by boiling the specimens in Immunosaver (Nissin EM, Tokyo, Japan) diluted 1:200 for 45 minutes at 98 °C. The sections were permeabilized with 0.1% Triton X-100 (Sigma-Aldrich) for 15 minutes. After blocking with Dako blocking reagent for 30 minutes, sections were incubated with primary antibodies overnight at 4°C in a humidified box. Sections were then incubated with secondary antibodies labeled with Alexa Fluor 488 and 594 (Invitrogen). Slides were mounted with VECTASHIELD mounting medium with DAPI (Vector Labs, CA, USA). Stained sections were imaged using a BZ-X710 microscope (Keyence, Osaka, Japan).

### Antibodies and reagents

Antibodies and reagents were as follows: anti-PD-L1 (rabbit, 1:1000, #ab205921, Abcam, Cambridge, UK); anti-TNC (rabbit, 1:100, #HPA004823, Sigma-Aldrich); anti-CCL2 (rabbit, 1:100, #HPA019163, Sigma-Aldrich); anti-RAGE (rabbit, 1:1000, #ab216329, Abcam); anti-SFTPC (rabbit, 1:1000, #HPA010928, Sigma-Aldrich); anti-ABCA3 (mouse, 1:1000, #WMAB-ABCA3-17, Seven Hills Bioreagents, OH, USA); anti-macrophage inflammatory protein 3 alpha (rabbit, 1:1000, #ab224188, Abcam); anti- CXCL1 (mouse, 1:100, #MAB275, R&D Systems, MN, USA); anti-CXCL8 (mouse, 1:100, #MAB208, R&D Systems); and Hoechst 33242 (1:1000, #H342, Sigma-Aldrich).

### Statistical Analysis

Student’s t-test was used for comparison of two data sets. Correlation coefficients were calculated by Spearman correlation. Analysis of variance (ANOVA) was used for multiple comparisons, followed by Tukey’s or Dunnett’s multiple comparison to find where the differences lie. Significance was defined as P < 0.05. The statistical software used was Prism version 8 (GraphPad Software, Inc., CA, USA).

## Author contributions

N.W ., J.N., Y.F. and Y.Y. conceived and designed the study. N.W. and Y.M. performed the experiments. N.W., J.N. and Y.Y. performed the data analysis. N.W., T.K., I.S., K.O., J.A., and K.K. interpreted the data. T.O. provided the patient samples. N.W., J.N., Y.F., J.A., and Y.Y. wrote the manuscript. Y.Y. supervised this project. All authors reviewed and edited the manuscript.

## Acknowledgements

We are grateful to Dr. S. Arimura for critically reading the manuscript and all members of the lab for stimulating discussions during the preparation of this manuscript. This publication is part of the Single Cell Medical Network of Japan. Schematics in figures were made using an academic license of Biorender software. This work was supported by Project for MEXT KAKENHI (Grant-in-Aid for Young Scientists (A); grant number: 17H04991, Grant-in-Aid for Scientific Research (B); grant number: 21H02721, Grant-in-Aid for JSPS Fellows: 20J01794), Research grant from The Naito Foundation, GSK Japan Research Grant, Mochida Memorial Foundation for Medical and Pharmaceutical Research, and Takeda Science Foundation.

## Supplemental material

**Table S1. Patient demographics and clinical data**

**Table S2. scRNA-seq information of each sample**

**Table S3. Number of cells per cluster per donor**

**Table S4. The average expression of the top 10 genes in each cluster**

**Table S5. Ratio of AT1 and AT2 cells to total cell count**

**Table S6. CCI score: Epithelial cells to Immune cells in COPD**

**Table S7. CCI score: Immune cells to Epithelial cells in COPD**

**Table S8. CCI score: Epithelial cells to Immune cells in Non-COPD smokers**

**Table S9. CCI score: Immune cells to Epithelial cells in Non-COPD smokers**

**Table S10. CCI score: Epithelial cells to Immune cells in Never smokers**

**Table S11. CCI score: Immune cells to Epithelial cells in Never smokers**

**Table S12. Statistical analysis of AT2 cluster variation**

---

**Supplemental Figure 1.**
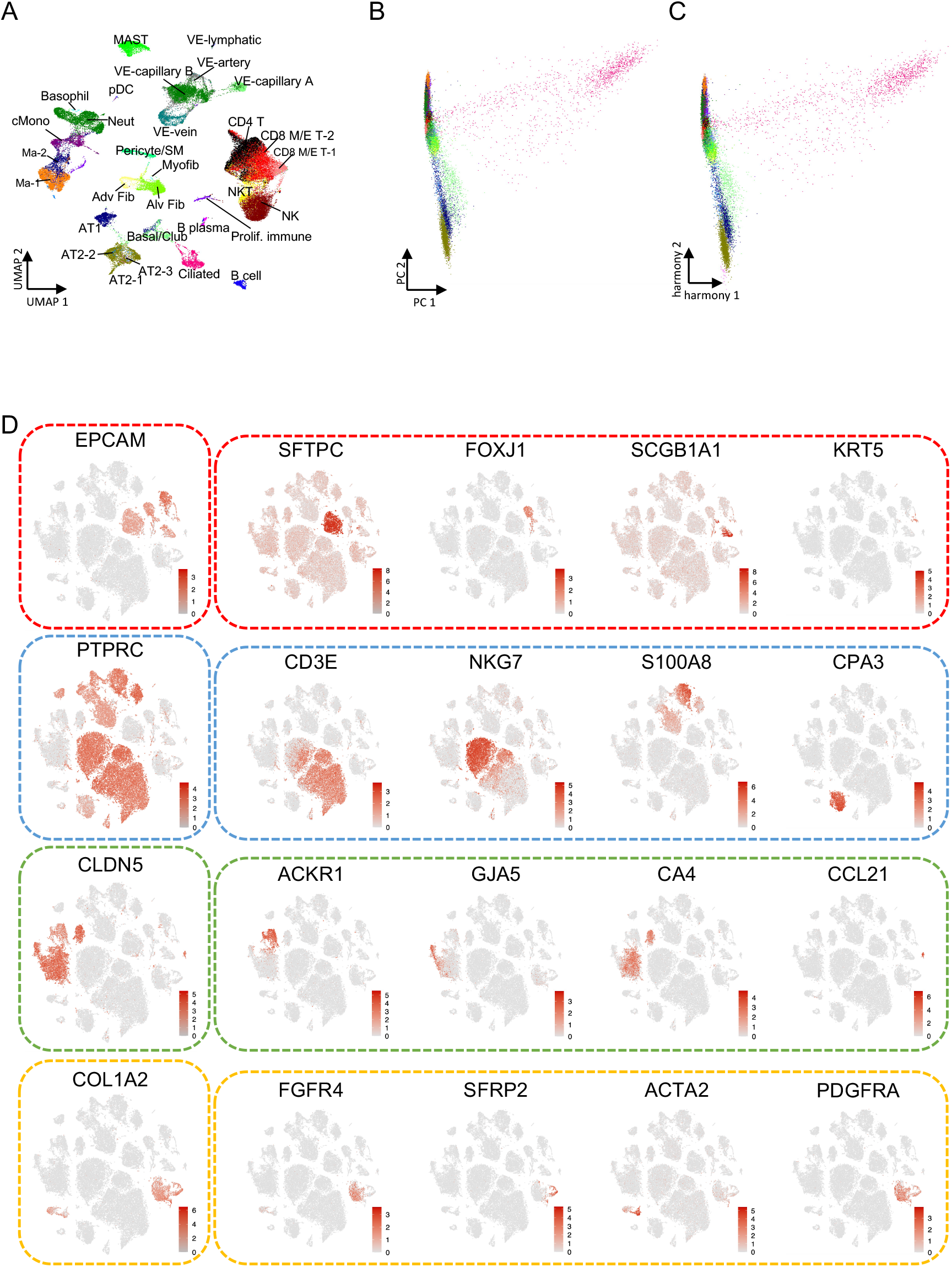
Unbiased clustering of human lung tissue cells from COPD patients, non-COPD smokers, and never-smokers. **A.** a UMAP plot with human lung tissue cells for unbiased clustering. **B.** PCA mapping with human lung tissue cells. **C.** PCA mapping with reduced batch effect using “Harmony”. **D.** t-SNE showing the expression of individual genes used for cell-type assignment of different cell subsets, such as epithelial cells, immune cells, endothelial cells, and stromal cells. NK, natural killer cell; NKT, natural killer T cell; Ma, macrophage; Neut, neutrophil; cMono, classical monocyte; pDC, plasmacytoid dendritic cell; VE, vascular endothelial; AT, alveolar type; CD4 T, CD4^+^ T cell; CD8 M/E T, CD8^+^ memory/effector T cell; Alv Fib, alveolar fibroblast; Adv Fib, adventitial fibroblast; Myofib, myofibroblast; Proif.immune, proliferating immune cell; SM, smooth muscle.

**Supplemental Figure 2.**
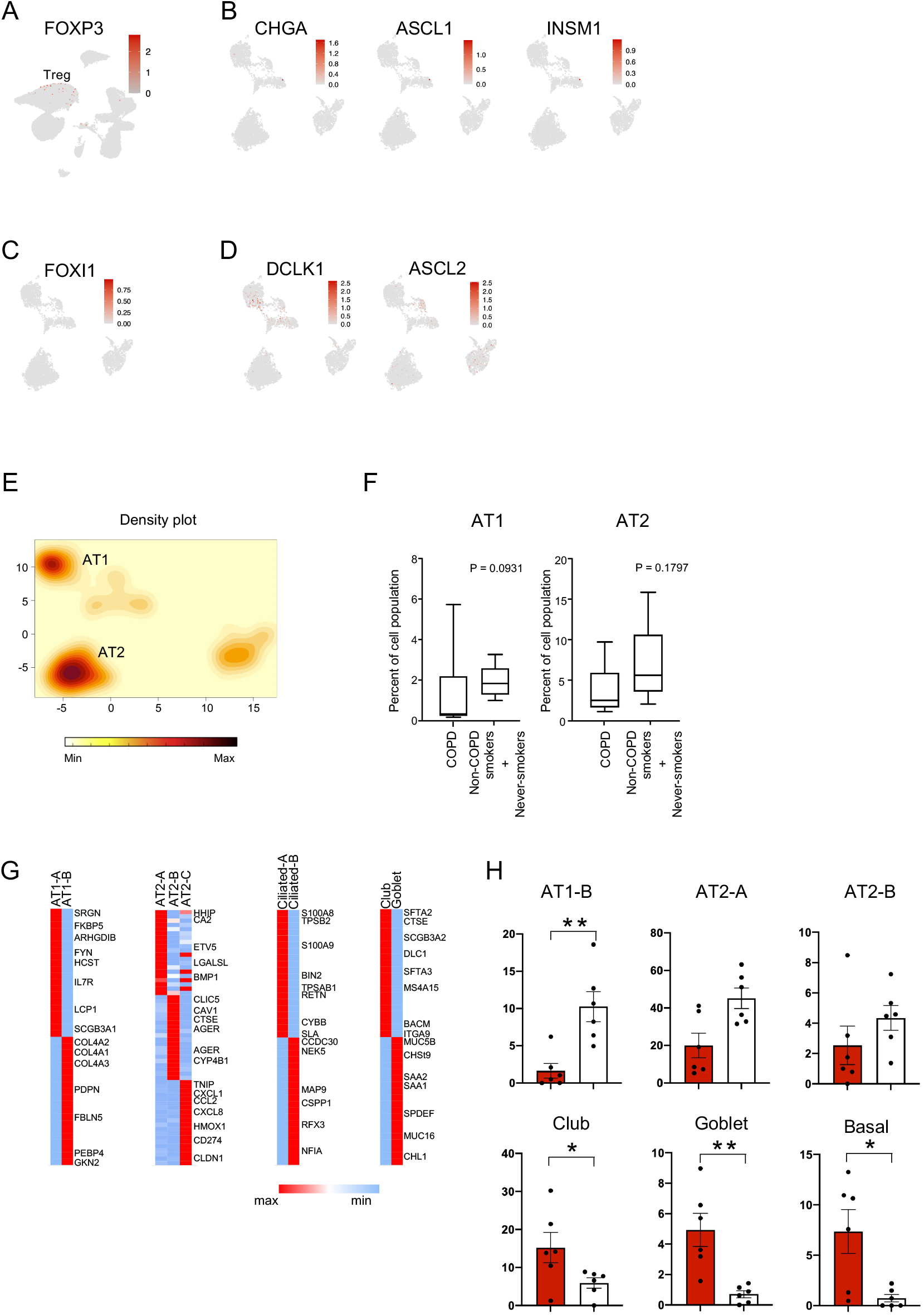
Expression profiles of specific epithelial marker genes. **A.** a UMAP plot of Foxp3 for depicting Tregs in the CD4 T cluster. **B.** UMAP plots of CHGA, ASCL1, and INSM1 for depicting neuroendocrine cells. **C.** a UMAP plot of FOXI1 for depicting ionocytes. **D.** UMAP plots of DCLK1 and ASCL2 for depicting tuft cells. **E.** a density plot of epithelial components in human lung cells. **F.** Number of AT1 and AT2 cells in COPD and non-COPD (non-COPD smokers and never-smokers) patients. **G.** Heatmap of differentially expressed genes in each epithelial cluster. The average expression (log2[TPM + 1]) of the top 20 genes (row) in each epithelial cluster (columns) is shown. **H.** Bar charts displaying the percentages of subpopulations such as AT1-B, AT2-A, AT2-B, club, goblet, and basal clusters in COPD and non-COPD (non-COPD smokers and never-smokers) patients. **\***p < 0.05, **\*\***p < 0.01 by Mann-Whitney test.

**Supplemental Figure 3.**
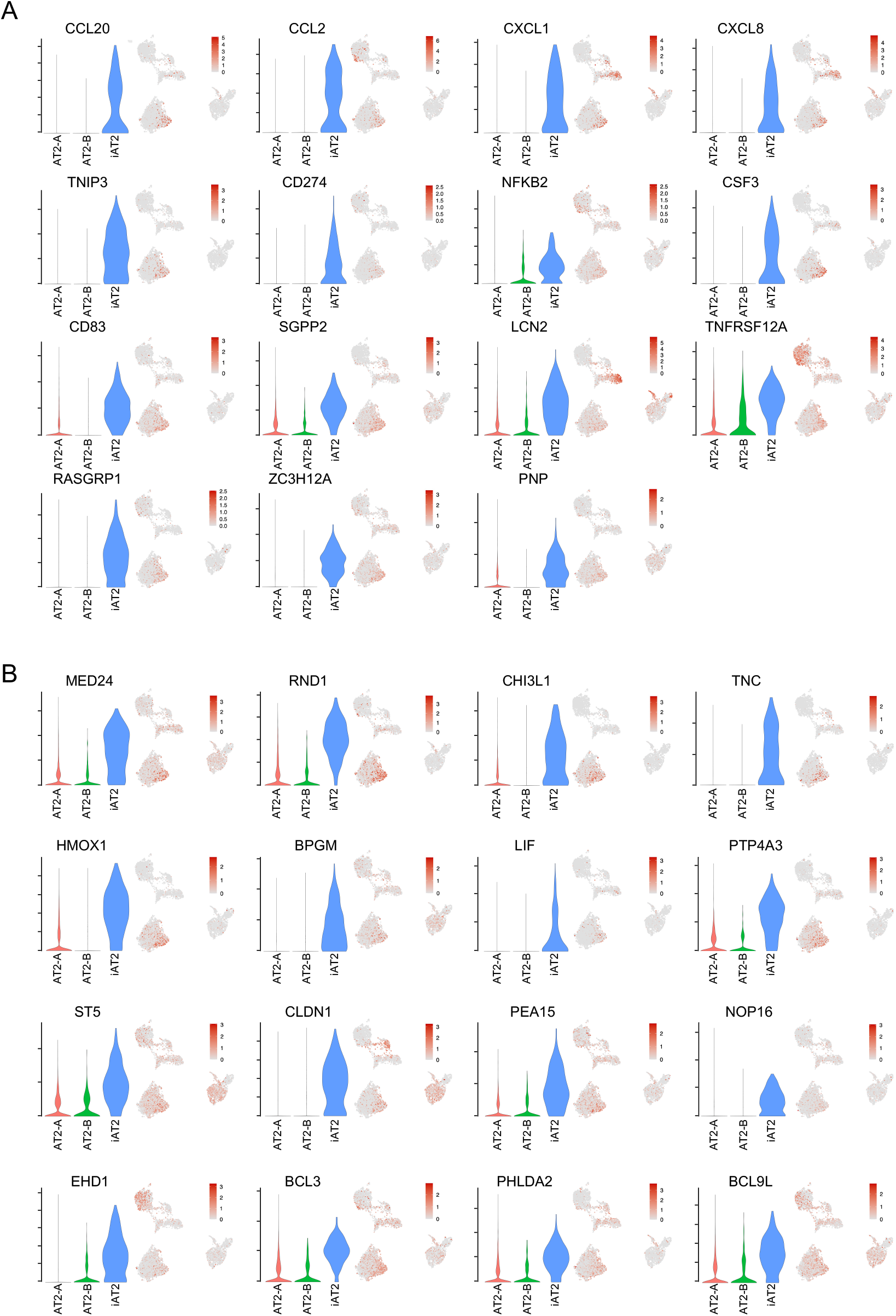
Expression profiles of identified inflammatory AT2 cluster markers. **A.** Violin and UMAP plots of inflammatory-related genes displaying the AT2- C (iAT2) cluster. **B.** Violin and UMAP plots of selectively expressed genes in the AT2-C (iAT2) cluster.

**Supplemental Figure 4.**
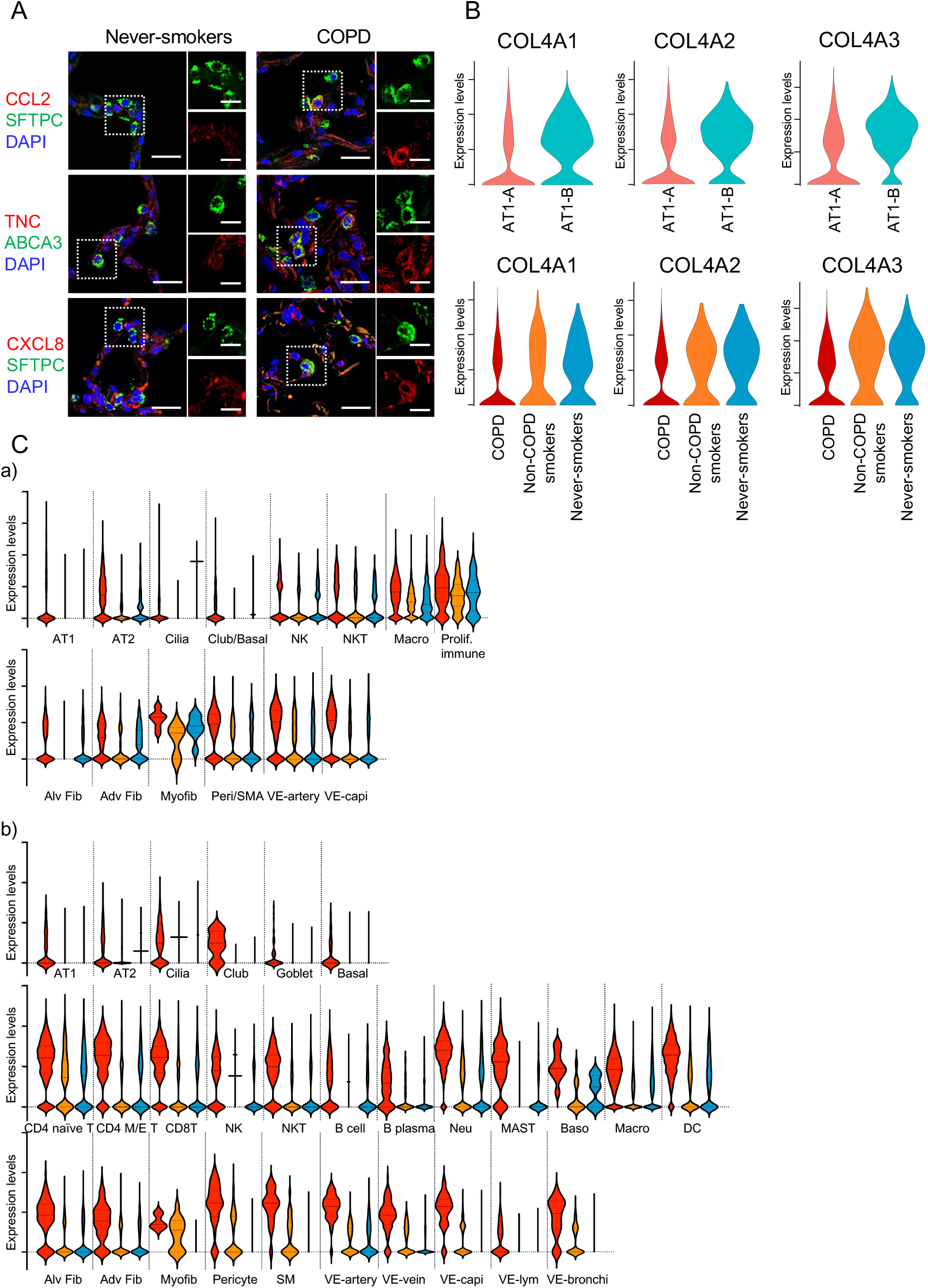
Confirmation of inflammatory responses and selected marker gene expressions. **A.** Representative images immunostained with CCL2, TNC, and CXCL8 with AT2 cell markers (SFTPC or ABCA3) in COPD and never-smoker lung sections. **B**. Violin plots for COL4A1, COL4A2, and COL4A3 in AT1 clusters. **C.** Violin plots for FKBP5 in selected cell types. a) present dataset; b) combined publicly-available datasets.

**Supplemental Figure 5.**
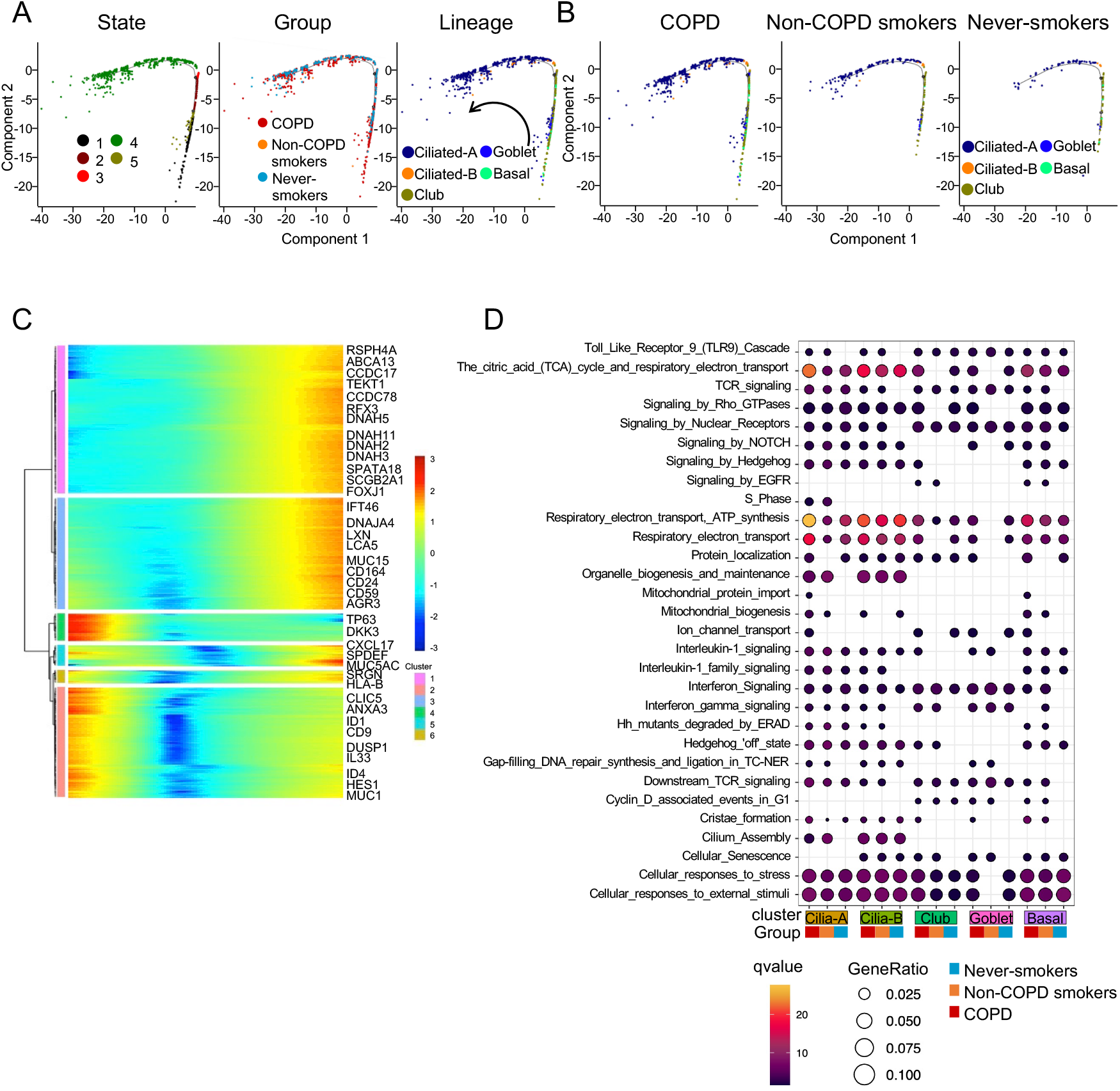
Pseudotime prediction of basal lineage differentiation. **A.** Pseudotime developmental trajectory analysis of basal cell lineages. Left panel: state, middle panel: group (COPD, non-COPD smokers, and never-smokers), right panel: basal clusters. Arrow: prediction of differentiation flow. **B.** Pseudotime time plots for COPD, non-COPD smokers, and never-smokers. **C.** Expression heatmap showing differentially expressed genes between two transition states (FDR < 0.01). Selected genes are highlighted. **D.** Pathway enrichment analysis of basal cell clusters among COPD, non- COPD smokers, and never-smokers. Color scale represents FDR q value. Circle size represents the likelihood ratio.

**Supplemental Figure 6.**
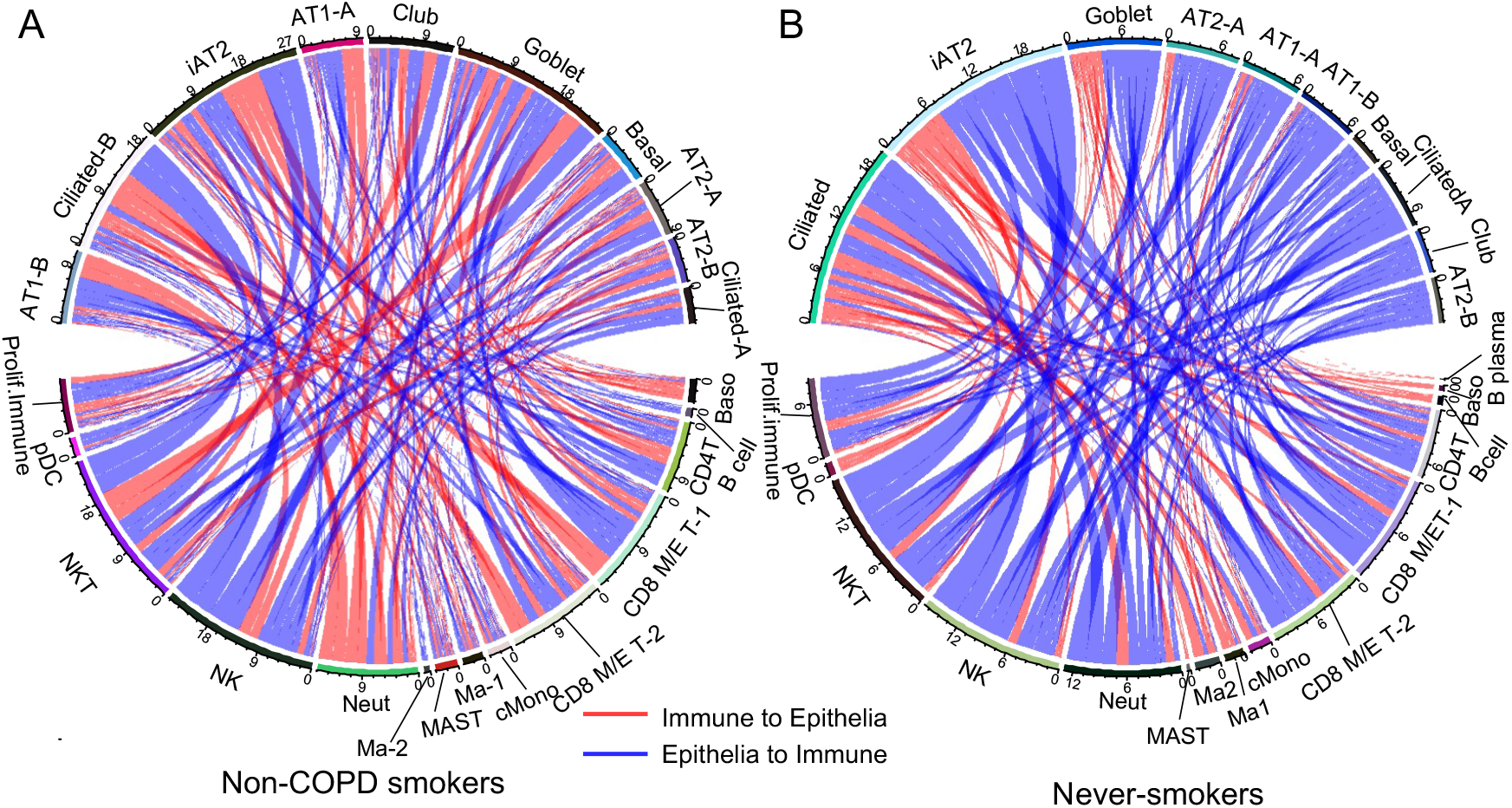
Inferred cell-to-cell interaction networks in non-COPD smokers and never-smokers. **A, B.** Circos plot of cell-to-cell interactions featuring inflammatory cytokines and chemokines in non-COPD smokers (**A**) and never-smokers (**B**). Red lines represent predicted immune cell-ligand to epithelial cell-receptor interactions. Blue lines represent predicted epithelial cell-ligand to immune cell-receptor interactions.

## Notes

### Competing Interest Statement

The authors have declared no competing interest.

### Author Declarations

Human lung tissue from never-smokers, non-COPD smokers, and patients with COPD was obtained from patients undergoing pneumonectomy or lobectomy for primary lung cancer. The diagnosis of COPD was established using the Global Initiative for Chronic Obstructive Lung Disease criteria. Informed consent was obtained from all surgical participants as part of an approved ongoing research protocol by the ethical committee of The Jikei University School of Medicine (#20-153 (5443)).

## References

1. Boucherat O, Morissette MC, Provencher S, Bonnet S, Maltais F. Bridging Lung Development with Chronic Obstructive Pulmonary Disease. Relevance of Developmental Pathways in Chronic Obstructive Pulmonary Disease Pathogenesis. Am J Respir Crit Care Med. Feb 15 2016;193(4):362–75. doi:10.1164/rccm.201508-1518PP

2. Crotty Alexander LE, Shin S, Hwang JH. Inflammatory Diseases of the Lung Induced by Conventional Cigarette Smoke: A Review. Chest. Nov 2015;148(5):1307–1322. doi:10.1378/chest.15-0409

3. Rabe KF, Watz H. Chronic obstructive pulmonary disease. Lancet. May 13 2017;389(10082):1931–1940. doi:10.1016/S0140-6736(17)31222-9

4. Tuder RM, Petrache I. Pathogenesis of chronic obstructive pulmonary disease. J Clin Invest. Aug 2012;122(8):2749–55. doi:10.1172/JCI60324

5. Beers MF, Moodley Y. When Is an Alveolar Type 2 Cell an Alveolar Type 2 Cell? A Conundrum for Lung Stem Cell Biology and Regenerative Medicine. Am J Respir Cell Mol Biol. Jul 2017;57(1):18–27. doi:10.1165/rcmb.2016-0426PS

6. Selman M, Pardo A. Revealing the pathogenic and aging-related mechanisms of the enigmatic idiopathic pulmonary fibrosis. an integral model. Am J Respir Crit Care Med. May 15 2014;189(10):1161–72. doi:10.1164/rccm.201312-2221PP

7. Garcia O, Hiatt MJ, Lundin A, et al. Targeted Type 2 Alveolar Cell Depletion. A Dynamic Functional Model for Lung Injury Repair. Am J Respir Cell Mol Biol. Mar 2016;54(3):319–30. doi:10.1165/rcmb.2014-0246OC

8. Randell SH. Airway epithelial stem cells and the pathophysiology of chronic obstructive pulmonary disease. Proc Am Thorac Soc. Nov 2006;3(8):718–25. doi:10.1513/pats.200605-117SF

9. Gamez AS, Gras D, Petit A, et al. Supplementing defect in club cell secretory protein attenuates airway inflammation in COPD. Chest. Jun 2015;147(6):1467–1476. doi:10.1378/chest.14-1174

10. Habermann AC, Gutierrez AJ, Bui LT, et al. Single-cell RNA sequencing reveals profibrotic roles of distinct epithelial and mesenchymal lineages in pulmonary fibrosis. Sci Adv. Jul 2020;6(28):eaba1972. doi:10.1126/sciadv.aba1972

11. Reyfman PA, Walter JM, Joshi N, et al. Single-Cell Transcriptomic Analysis of Human Lung Provides Insights into the Pathobiology of Pulmonary Fibrosis. Am J Respir Crit Care Med. Jun 15 2019;199(12):1517–1536. doi:10.1164/rccm.201712-2410OC

12. Adams TS, Schupp JC, Poli S, et al. Single-cell RNA-seq reveals ectopic and aberrant lung-resident cell populations in idiopathic pulmonary fibrosis. Sci Adv. Jul 2020;6(28):eaba1983. doi:10.1126/sciadv.aba1983

13. Vieira Braga FA, Kar G, Berg M, et al. A cellular census of human lungs identifies novel cell states in health and in asthma. Nat Med. Jul 2019;25(7):1153–1163. doi:10.1038/s41591-019-0468-5

14. Li X, Noell G, Tabib T, et al. Single cell RNA sequencing identifies IGFBP5 and QKI as ciliated epithelial cell genes associated with severe COPD. Respir Res. Apr 6 2021;22(1):100. doi:10.1186/s12931-021-01675-2

15. Korsunsky I, Millard N, Fan J, et al. Fast, sensitive and accurate integration of single-cell data with Harmony. Nat Methods. Dec 2019;16(12):1289–1296. doi:10.1038/s41592-019-0619-0

16. Tran HTN, Ang KS, Chevrier M, et al. A benchmark of batch-effect correction methods for single-cell RNA sequencing data. Genome Biol. Jan 16 2020;21(1):12. doi:10.1186/s13059-019-1850-9

17. Travaglini KJ, Nabhan AN, Penland L, et al. A molecular cell atlas of the human lung from single-cell RNA sequencing. Nature. Nov 2020;587(7835):619–625. doi:10.1038/s41586-020-2922-4

18. Jiang S, Park DW, Tadie JM, et al. Human resistin promotes neutrophil proinflammatory activation and neutrophil extracellular trap formation and increases severity of acute lung injury. J Immunol. May 15 2014;192(10):4795–803. doi:10.4049/jimmunol.1302764

19. Ezzie ME, Crawford M, Cho JH, et al. Gene expression networks in COPD: microRNA and mRNA regulation. Thorax. Feb 2012;67(2):122–31. doi:10.1136/thoraxjnl-2011-200089

20. Randazzo PA, Andrade J, Miura K, et al. The Arf GTPase-activating protein ASAP1 regulates the actin cytoskeleton. Proc Natl Acad Sci U S A. Apr 11 2000;97(8):4011–6. doi:10.1073/pnas.070552297

21. Knight DA, Holgate ST. The airway epithelium: structural and functional properties in health and disease. Respirology. Dec 2003;8(4):432–46. doi:10.1046/j.1440-1843.2003.00493.x

22. Plasschaert LW, Zilionis R, Choo-Wing R, et al. A single-cell atlas of the airway epithelium reveals the CFTR-rich pulmonary ionocyte. Nature. Aug 2018;560(7718):377–381. doi:10.1038/s41586-018-0394-6

23. Howitt MR, Lavoie S, Michaud M, et al. Tuft cells, taste-chemosensory cells, orchestrate parasite type 2 immunity in the gut. Science. Mar 18 2016;351(6279):1329–33. doi:10.1126/science.aaf1648

24. Nabhan AN, Brownfield DG, Harbury PB, Krasnow MA, Desai TJ. Single-cell Wnt signaling niches maintain stemness of alveolar type 2 cells. Science. Mar 9 2018;359(6380):1118–1123. doi:10.1126/science.aam6603

25. Zacharias WJ, Frank DB, Zepp JA, et al. Regeneration of the lung alveolus by an evolutionarily conserved epithelial progenitor. Nature. Mar 8 2018;555(7695):251–255. doi:10.1038/nature25786

26. Villasenor-Altamirano AB, Moretto M, Maldonado M, et al. PulmonDB: a curated lung disease gene expression database. Sci Rep. Jan 16 2020;10(1):514. doi:10.1038/s41598-019-56339-5

27. Treekitkarnmongkol W, Hassane M, Sinjab A, et al. Augmented Lipocalin-2 Is Associated with Chronic Obstructive Pulmonary Disease and Counteracts Lung Adenocarcinoma Development. Am J Respir Crit Care Med. Jan 1 2021;203(1):90–101. doi:10.1164/rccm.202004-1079OC

28. Russo P, Tomino C, Santoro A, et al. FKBP5 rs4713916: A Potential Genetic Predictor of Interindividual Different Response to Inhaled Corticosteroids in Patients with Chronic Obstructive Pulmonary Disease in a Real-Life Setting. Int J Mol Sci. Apr 24 2019;20(8)doi:10.3390/ijms20082024

29. Trapnell C, Cacchiarelli D, Grimsby J, et al. The dynamics and regulators of cell fate decisions are revealed by pseudotemporal ordering of single cells. Nat Biotechnol. Apr 2014;32(4):381–386. doi:10.1038/nbt.2859

30. Yu G, He QY. ReactomePA: an R/Bioconductor package for reactome pathway analysis and visualization. Mol Biosyst. Feb 2016;12(2):477–9. doi:10.1039/c5mb00663e

31. Brightling C, Greening N. Airway inflammation in COPD: progress to precision medicine. Eur Respir J. Aug 2019;54(2)doi:10.1183/13993003.00651-2019

32. Rock JR, Gao X, Xue Y, Randell SH, Kong YY, Hogan BL. Notch-dependent differentiation of adult airway basal stem cells. Cell Stem Cell. Jun 3 2011;8(6):639–48. doi:10.1016/j.stem.2011.04.003

33. Shi W, Chen F, Cardoso WV. Mechanisms of lung development: contribution to adult lung disease and relevance to chronic obstructive pulmonary disease. Proc Am Thorac Soc. Dec 1 2009;6(7):558–63. doi:10.1513/pats.200905-031RM

34. Saito N, Araya J, Ito S, et al. Involvement of Lamin B1 Reduction in Accelerated Cellular Senescence during Chronic Obstructive Pulmonary Disease Pathogenesis. J Immunol. Mar 1 2019;202(5):1428–1440. doi:10.4049/jimmunol.1801293

35. Tsuji T, Aoshiba K, Nagai A. Alveolar cell senescence in patients with pulmonary emphysema. Am J Respir Crit Care Med. Oct 15 2006;174(8):886–93. doi:10.1164/rccm.200509-1374OC

36. Takasaka N, Araya J, Hara H, et al. Autophagy induction by SIRT6 through attenuation of insulin-like growth factor signaling is involved in the regulation of human bronchial epithelial cell senescence. J Immunol. Feb 1 2014;192(3):958–68. doi:10.4049/jimmunol.1302341

37. Ito S, Araya J, Kurita Y, et al. PARK2-mediated mitophagy is involved in regulation of HBEC senescence in COPD pathogenesis. Autophagy. 2015;11(3):547–59. doi:10.1080/15548627.2015.1017190

38. Demedts IK, Demoor T, Bracke KR, Joos GF, Brusselle GG. Role of apoptosis in the pathogenesis of COPD and pulmonary emphysema. Respir Res. Mar 30 2006;7:53. doi:10.1186/1465-9921-7-53

39. Mizumura K, Cloonan SM, Nakahira K, et al. Mitophagy-dependent necroptosis contributes to the pathogenesis of COPD. J Clin Invest. Sep 2014;124(9):3987–4003. doi:10.1172/JCI74985

40. Yoshida M, Minagawa S, Araya J, et al. Involvement of cigarette smoke-induced epithelial cell ferroptosis in COPD pathogenesis. Nat Commun. Jul 17 2019;10(1):3145. doi:10.1038/s41467-019-10991-7

41. Sauler M, McDonough JE, Adams TS, et al. [Preprint] https://doi.org/10.1101/2020.09.13.20193417. Posted on medRxiv September 4, 2020.

42. Plosa EJ, Benjamin JT, Sucre JM, et al. beta1 Integrin regulates adult lung alveolar epithelial cell inflammation. JCI Insight. Jan 30 2020;5(2)doi:10.1172/jci.insight.129259

43. Ahmadvand N, Khosravi F, Lingampally A, et al. Identification of a novel subset of alveolar type 2 cells enriched in PD-L1 and expanded following pneumonectomy. Eur Respir J. Apr 16 2021;doi:10.1183/13993003.04168-2020

44. Stuart T, Butler A, Hoffman P, et al. Comprehensive Integration of Single-Cell Data. Cell. Jun 13 2019;177(7):1888–1902 e21. doi:10.1016/j.cell.2019.05.031

45. Yu G, Wang LG, Han Y, He QY. clusterProfiler: an R package for comparing biological themes among gene clusters. OMICS. May 2012;16(5):284–7. doi:10.1089/omi.2011.0118

46. Nakayama J, Ito E, Fujimoto J, Watanabe S, Semba K. Comparative analysis of gene regulatory networks of highly metastatic breast cancer cells established by orthotopic transplantation and intra-circulation injection. Int J Oncol. Feb 2017;50(2):497–504. doi:10.3892/ijo.2016.3809

47. Saito R, Smoot ME, Ono K, et al. A travel guide to Cytoscape plugins. Nat Methods. Nov 2012;9(11):1069–76. doi:10.1038/nmeth.2212

48. Shannon P, Markiel A, Ozier O, et al. Cytoscape: a software environment for integrated models of biomolecular interaction networks. Genome Res. Nov 2003;13(11):2498–504. doi:10.1101/gr.1239303

49. Gu P, Chen H. Modern bioinformatics meets traditional Chinese medicine. Brief Bioinform. Nov 2014;15(6):984–1003. doi:10.1093/bib/bbt063

